# International attitudes on COVID-19 vaccination: repeat national cross-sectional surveys across 15 countries

**DOI:** 10.1101/2021.03.08.21252449

**Authors:** Roberto Fernandez Crespo, Manar Shafat, Natalie Melas-Kyriazi, Lisa Gould, Sarah Jones, Ana Luisa Neves, Melanie S Leis, Hendramoorthy Maheswaran, Ara Darzi

## Abstract

**Objective:** To examine the general public’s views around willingness to receive COVID-19 vaccines and concerns regarding their safety.

**Design:** Repeat cross-sectional surveys.

**Setting:** Online surveys in Australia, Canada, Denmark, Finland, France, Germany, Italy, Japan, Netherlands, Norway, Singapore, South Korea, Spain, Sweden and the United Kingdom

**Participants:** National samples of adults aged >=18 years in November 2020 and January 2021.

**Main outcomes measures:** The proportion of adults reporting: willingness to receive a COVID-19 vaccination; concern regarding side-effects from vaccinations; concerns over contraction COVID-19, and beliefs around vaccine provision in their country. Changes between the November and January surveys are also reported.

**Results:** Across the 15 countries, the proportion of respondents reporting they would have the COVID-19 vaccine increased from 40.7% (range: 25.0-55.1) to 55.2% (range: 34.8-77.5), proportion reporting worried about the side-effects of vaccine decreased from 53.3% (range: 42.1-66.7) to 47.9% (range: 28.0-66.1). On the second survey, willingness to receive vaccine remained low in females (49.4%, range: 30.2-79.1), aged 18-39 years (42.1%, range: 25.9-71.7), those not working or unemployed (48.9, range: 18.8-67.0), students (45.9%, range: 22.8-70.0), and those with children at home (46.5%, range: 32.4-68.9). Concerns regarding safety of vaccine remained high in females (53.7%, range: 31.8-70.4), aged 18-39 years (50.8%, range: 28.2-60.7), aged 40-64 years (51.3%, range: 30.7-68.5), those working (50.5%, range: 26.7-65.0), those not working or unemployed (53.3, range: 35.4-73.8) and those with children at home (55.8%, range: 36.5-64.7).

**Conclusion:** COVID-19 vaccine hesitancy decreased considerably over a relatively short time coinciding with the discovery of effective vaccines. The public remain concerned about their safety, and public health messaging will need to emphasis their safety especially amongst females, parents and younger adults.

## Introduction

The discovery of multiple effective vaccines against COVID-19 offers a path out of the global pandemic that has already taken over 2 million lives. Success of global vaccination will depend on achieving herd immunity, from individuals acquiring immunity through vaccination or after infection. For COVID-19 this may require more than three thirds of populations receiving an effective vaccine^1^. However, there are concerns that many individuals are reluctant to get a COVID-19 vaccine, with reasons ranging from concerns regarding their safety to perceptions of a low personal risk of severe illness^2-4^. Studies undertaken prior to the discovery of effective vaccines have found up to a third of the population may be resistant or hesitant to COVID-19 vaccination^2 5 6^.

In this study we sought to examine people’s willingness to receive the COVID-19 vaccine and whether vaccine hesitancy had reduced after effective vaccines were discovered and provided in many countries. Additionally, we explored respondents’ perceptions about vaccine safety, and their views on whether their government would be able to deliver mass vaccination. We were able to compare changes in attitudes in this data from November 2020 to January 2021. We use repeated cross-sectional nationally representative data collected from 15 countries; and explored how these views differ and have changed across different socio-demographic groups and higher risk individuals.

## Methods

### Participants and procedures

This study uses two waves of nationally representative data collected by YouGov in 15 countries (Australia; Canada; Denmark; Finland; France; Germany; Italy; Japan; Netherlands; Norway; Singapore; South Korea; Spain; Sweden; United Kingdom). These cross-sectional surveys were started in April 2020 to capture health behaviour and public perceptions around COVID-19 and are repeated every two weeks. For each country and survey, YouGov samples a nationally representative population to complete surveys online, from a larger panel of respondents in each country. YouGov generates survey weights to adjust the samples to be nationally representative.

The survey consists of a core set of demographic questions and additional questions relating to COVID-19 and preventative measures to curtail the pandemic. Since November 2020, respondents have been asked about their views on COVID-19 vaccination. This study is based on data collected when these questions were introduced (9-15^th^ November 2020) and the most recent survey undertaken in each country (11^th^ January 2021 to 31^st^ January 2021).

### Measures

The main outcome measures included in this analysis related to respondents’ responses to four COVID-19 related questions. The questions asked respondents to what extent they agreed or disagreed with the statements; (1) *I am worried about getting COVID-19*; (2) *If a COVID-19 vaccine were made available to me this week, I would definitely get it*; (3) *I am worried about potential side effects of a COVID-19 vaccine*; (4) *I believe government health authorities in my country will provide me with an effective COVID-19 vaccine*. Respondents were asked to indicate their response on a scale between 1 (strongly agreed) and 5 (strongly disagreed).

Sex, age and employment status were reported by survey respondents. Respondents were asked whether they had any one of the following health conditions: Arthritis; Asthma; Cancer; Cystic fibrosis; Chronic obstructive pulmonary disease; Diabetes; Epilepsy; Heart disease; High blood pressure; High cholesterol; HIV/AIDS; Mental health condition; Multiple Sclerosis. Respondents who responded ‘yes’ to any of these conditions were classified as having long-term health condition. Additional categories for this variable included: No; prefer not to say. Respondents were asked if they lived with children under the age of 18.

Respondents were asked whether they worked outdoors (construction; delivering to homes; food retail; healthcare; logistics or other transportation; manufacturing; policing or prisons; public transport; school; social care) in the week of the survey. Responses were re-categorised as whether or not respondent was a frontline worker (yes; no; other/not sure).

### Data analysis

We analysed the data for each country separately and used the survey weights to account for differential probabilities for selection to the survey. Responses to the four questions on Covid-19 were recoded as a dichotomous response of agree (strongly agree/1; or 2) and not agree (strongly disagreed/5; 4; or 3). We present descriptive analyses (percentage agree) for the whole country, and by sex, age group (18-39 years; 40-65 years; >=65 years); employment status (working; retired; student; not working/unemployed); have children at home (yes; no prefer not to say); long-term health condition (yes; no; prefer not to say), and frontline worker (yes; no; other/not sure).

We present the mean percentage change and 95% confidence interval within each country, between responses in the first wave of surveys (9-15^th^ November 2020) and the second wave of surveys (11^th^-31^st^ January 2021). We classified this change in percentage as increased (>5%; 0-5%); or decreased (>5%; 0-5%). We also present the median and low/high percentage of respondents who agreed, across the sample of 15 countries on each survey date.

## Results

Table 1 shows the survey dates for the 15 countries and number of respondents sampled on each occasion. For all 15 countries the first surveys were undertaken between 9^th^ and 15^th^ November 2020 and the second surveys undertaken between 11^th^ and 31^st^ January 2021. The number of respondents sampled in each country, on each survey, ranged from 483 to 1032. 12/15 countries had commenced their COVID-19 vaccination programmes prior to the second survey.

**Table 1:** Description of surveys and countries.

| Country | Survey date | Number of Respondents | Vaccination doses administered per 100 people on date of 2<sup>nd</sup> survey* |
| --- | --- | --- | --- |
| Australia | 9<sup>th</sup> Nov to 15<sup>th</sup> Nov 2020<br>11<sup>th</sup> Jan to 17<sup>th</sup> Jan 2021 | 1001<br>1001 | Not started |
| Canada | 9<sup>th</sup> Nov to 15<sup>th</sup> Nov 2020<br>18<sup>th</sup> Jan to 24<sup>th</sup> Jan 2021 | 1003<br>968 | 1.12 |
| Denmark | 9<sup>th</sup> Nov to 15<sup>th</sup> Nov 2020<br>11<sup>th</sup> Jan to 17<sup>th</sup> Jan 2021 | 1006<br>1000 | 2.03 |
| Finland | 9<sup>th</sup> Nov to 15<sup>th</sup> Nov 2020<br>18<sup>th</sup> Jan to 24<sup>th</sup> Jan 2021 | 1001<br>1006 | 0.98 |
| France | 9<sup>th</sup> Nov to 15<sup>th</sup> Nov 2020<br>18<sup>th</sup> Jan to 24<sup>th</sup> Jan 2021 | 1000<br>1032 | 0.69 |
| Germany | 9<sup>th</sup> Nov to 15<sup>th</sup> Nov 2020<br>11<sup>th</sup> Jan to 17<sup>th</sup> Jan 2021 | 1000<br>1000 | 0.85 |
| Italy | 9<sup>th</sup> Nov to 15<sup>th</sup> Nov 2020<br>25<sup>th</sup> Jan to 31<sup>st</sup> Jan 2021 | 1005<br>1004 | 2.5 |
| Japan | 9<sup>th</sup> Nov to 15<sup>th</sup> Nov 2020<br>25<sup>th</sup> Jan to 31<sup>st</sup> Jan 2021 | 505<br>499 | Not started |
| Netherlands | 9<sup>th</sup> Nov to 15<sup>th</sup> Nov 2020<br>25<sup>th</sup> Jan to 31<sup>st</sup> Jan 2021 | 509<br>504 | 0.86 |
| Norway | 9<sup>th</sup> Nov to 15<sup>th</sup> Nov 2020<br>25<sup>th</sup> Jan to 31<sup>st</sup> Jan 2021 | 1001<br>1002 | 1.51 |
| Singapore | 9<sup>th</sup> Nov to 15<sup>th</sup> Nov 2020<br>11<sup>th</sup> Jan to 17<sup>th</sup> Jan 2021 | 1000<br>1001 | 0.06 |
| South Korea | 9<sup>th</sup> Nov to 15<sup>th</sup> Nov 2020<br>18<sup>th</sup> Jan to 24<sup>th</sup> Jan 2021 | 499<br>483 | Not started |
| Spain | 9<sup>th</sup> Nov to 15<sup>th</sup> Nov 2020<br>11<sup>th</sup> Jan to 17<sup>th</sup> Jan 2021 | 1009<br>1001 | 0.87 |
| Sweden | 9<sup>th</sup> Nov to 15<sup>th</sup> Nov 2020<br>25<sup>th</sup> Jan to 31<sup>st</sup> Jan 2021 | 1001<br>1000 | 2.25 |
| United Kingdom | 9<sup>th</sup> Nov to 15<sup>th</sup> Nov 2020<br>25<sup>th</sup> Jan to 31<sup>st</sup> Jan 2021 | 1005<br>1001 | 10.79 |

\*Data extracted from 'Our World in Data'

Table 2 displays the proportion of respondents in each country that agreed with each of the four questions on COVID-19, and the percentage change between the two surveys. Table 3 displays the median estimate for the sample of 15 countries, for the whole population and across different population sub-groups. More detailed country-level data is provided in the Appendix. In 10/15 countries there was a significant increase in the proportion of respondents reporting they were worried about getting COVID-19 between the two surveys (Table 2), for 3/15 countries this increase was at least 5% (Figure 1). Across the 15 countries the median proportion of respondents that reported being worried about getting COVID-19 in the first and second surveys was 41.1% (range: 28.7-67.3) and 41.8% (range: 34.6-69.9), respectively (Table 3).

**Table 2:** Percentage ‘agreeing’ with COVID-19 related questions in each of the 15 countries.

| Country | % Would vaccinate |  |  | % Worried about side effects |  |  | % Believe government health authority will provide |  |  | % Worried about getting COVID-19 |  |  |
| --- | --- | --- | --- | --- | --- | --- | --- | --- | --- | --- | --- | --- |
|  | Wave 1 | Wave 2 | Change + 95%CI | Wave 1 | Wave 2 | Change + 95%CI | Wave 1 | Wave 2 | Change + 95%CI | Wave 1 | Wave 2 | Change + 95%CI |
| Australia | 55.1 | 48.5 | -6.6 (-6.6, -6.6) | 47.1 | 50.2 | 3.1 (3.0, 3.2) | 60.2 | 55.0 | -5.2 (-5.2, -5.3) | 36.8 | 36.2 | -0.6 (-0.6, -0.6) |
| Canada | 44.2 | 55.2 | 11.0 (10.9, 11.2) | 53.3 | 47.9 | -5.4 (-5.4, -5.5) | 49.4 | 58.5 | 9.1 (9.0, 9.2) | 38.5 | 42.4 | 3.9 (3.8,4.0) |
| Denmark | 53.7 | 67.0 | 13.3 (13.1, 13.5) | 42.1 | 35.4 | -6.7 (-6.7, -6.7) | 63.5 | 72.6 | 9.1 (9.0, 9.2) | 37.5 | 39.6 | 2.1 (2.0, 2.2) |
| Finland | 36.6 | 57.0 | 20.4 (20.2, 20.7) | 57.3 | 47.2 | -10.1 (-10.2, -10.0) | 47.3 | 53.8 | 6.5 (6.4, 6.6) | 34.0 | 35.1 | 1.1 (1.1, 1.2) |
| France | 25.0 | 39.2 | 14.2 (14.0, 14.4) | 66.2 | 55.2 | -11 (-11.1, -10.9) | 36.3 | 38.1 | 1.8 (1.7, 1.9) | 43.1 | 41.8 | -1.3 (-1.3, -1.3) |
| Germany | 40.0 | 53.0 | 13.0 (12.8, 13.2) | 51.7 | 50.8 | -0.9 (-0.9, -0.9) | 56.1 | 57.3 | 1.2 (1.1, 1.3) | 37.9 | 38.4 | 0.5 (0.5, 0.6) |
| Italy | 40.7 | 56.1 | 15.4 (15.2, 15.6) | 48.4 | 40.8 | -7.6 (-7.6, -7.6) | 44.2 | 52.5 | 8.3 (8.2, 8.4) | 46.8 | 45.2 | -1.6 (-1.6, -1.6) |
| Japan | 39.1 | 39.9 | 0.8 (0.7, 0.9) | 64.7 | 66.1 | 1.4 (1.3, 1.5) | 43.6 | 48.7 | 5.1 (5.0, 5.2) | 67.3 | 69.9 | 2.6 (2.5, 2.7) |
| Netherlands | 44.6 | 63.1 | 18.5 (18.3, 18.8) | 46.6 | 41.3 | -5.3 (-5.3, -5.3) | 47.3 | 56.6 | 9.3 (9.1, 9.5) | 28.7 | 34.6 | 5.9 (5.8, 6.0) |
| Norway | 41.9 | 56.5 | 14.6 (14.4, 14.8) | 49.9 | 44.0 | -5.9 (-5.9, -5.9) | 54.1 | 57.0 | 2.9 (2.8, 3.0) | 41.1 | 40.7 | -0.4 (-0.4, -0.4) |
| Singapore | 36.1 | 34.8 | -1.3 (-1.3, -1.3) | 58.9 | 63.6 | 4.7 (4.6, 4.8) | 64.2 | 60.3 | -3.9 (-3.9, -3.9) | 47.8 | 49.2 | 1.4 (1.3, 1.5) |
| South Korea | 48.7 | 43.7 | -5.0 (-5.0, -5.0) | 55.7 | 57.6 | 1.9 (1.8, 2.0) | 54.7 | 53.6 | -1.1 (-1.2, -1.1) | 55.1 | 58.3 | 3.2 (3.1, 3.3) |
| Spain | 28.0 | 52.1 | 24.1 (23.8, 24.4) | 66.7 | 57.4 | -9.3 (-9.4, -9.3) | 41.0 | 54.8 | 13.8 (13.6, 14.0) | 60.1 | 66.0 | 5.9 (5.8, 6.0) |
| Sweden | 34.9 | 57.6 | 22.7 (22.4, 23.0) | 54.4 | 41.2 | -13.2 (-13.3, -13.1) | 43.0 | 50.6 | 7.6 (7.5, 7.7) | 40.0 | 38.3 | -1.7 (-1.7, -1.7) |
| United Kingdom | 54.3 | 77.5 | 23.2 (22.9, 23.5) | 46.3 | 28.0 | -18.3 (-18.4, -18.2) | 50.7 | 70.5 | 19.8 (19.6, 20.0) | 44.2 | 50.1 | 5.9 (5.8, 6.0) |

**Table 3:** Percentage ‘agreeing’ with COVID-19 related questions across population subgroups*.

| Population characteristics |  | % Would vaccinate |  | % Worried about side effects |  | % Believe government health authority will provide |  | % Worried about getting COVID-19 |  |
| --- | --- | --- | --- | --- | --- | --- | --- | --- | --- |
|  |  | Wave 1<br>(median &<br>low/high) | Wave 2<br>(median &<br>low/high) | Wave 1<br>(median &<br>low/high) | Wave 2<br>(median &<br>low/high) | Wave 1<br>(median &<br>low/high) | Wave 2<br>(median &<br>low/high) | Wave 1<br>(median &<br>low/high) | Wave 2<br>(median &<br>low/high) |
| **All** |  | 40.7 (25.0, 55.1) | 55.2 (34.8, 77.5) | 53.3 (42.1, 66.7) | 47.9 (28.0, 66.1) | 49.4 (36.3, 64.2) | 55.0 (38.1, 72.6) | 41.1 (28.7, 67.3) | 41.8 (34.6, 69.9) |
| **Sex** |  |  |  |  |  |  |  |  |  |
|  | Male | 46.5 (32.8, 60.5) | 55.5 (39.2, 75.9) | 44.8 (29.6, 59.3) | 42.2 (24.1, 61.5) | 51.3 (42.3, 63.7) | 56.4 (42.7, 70.8) | 37.6 (23.8, 62.4) | 38.7 (28.7, 68.6) |
|  | Female | 35.0 (17.6, 50.0) | 49.4 (30.2, 79.1) | 58.4 (52.2, 74.6) | 53.7 (31.8, 70.4) | 47.3 (30.0, 64.6) | 54.2 (34.0, 74.3) | 45.4 (33.4, 71.9) | 46.1 (35.5, 71.1) |
| **Age group (years)** |  |  |  |  |  |  |  |  |  |
|  | 18-39 | 33.6 (19.5, 49.6) | 42.1 (25.9, 71.7) | 50.0 (42.1, 66.4) | 50.8 (28.1, 60.7) | 43.0 (29.3, 65.6) | 49.8 (30.1, 64.8) | 40.1 (24.8, 62.3) | 36.1 (27.8, 61.4) |
|  | 40-64 | 40.8 (23.0, 58.3) | 55.1 (35.6, 76.0) | 55.4 (44.7, 67.8) | 51.3 (30.7, 68.5) | 49.6 (37.6, 63.5) | 55.5 (39.7, 74.2) | 42.0 (31.3, 67.3) | 42.6 (31.2, 71.6) |
|  | 65+ | 54.8 (34.8, 71.9) | 68.8 (50.6, 90.0) | 50.8 (36.9, 73.1) | 39.4 (23.4, 74.9) | 62.7 (43.2, 80.3) | 68.3 (47.5, 87.3) | 45.2 (24.7, 75.4) | 49.7 (35.0, 79.4) |
| **Employment status** |  |  |  |  |  |  |  |  |  |
|  | Working | 41.7 (19.5, 52.9) | 51.1 (34.4, 76.7) | 53.3 (42.4, 66.9) | 50.5 (26.7, 65.0) | 45.7 (34.1, 62.9) | 54.2 (34.3, 69.7) | 41.6 (26.7, 68.9) | 42.7 (33.9, 69.9) |
|  | Student | 38.8 (25.5, 52.1) | 45.9 (22.8, 70.0) | 50.3 (33.4, 59.9) | 48.0 (13.1, 70.7) | 51.1 (35.3, 70.8) | 54.7 (45.4, 74.3) | 41.1 (27.2, 53.7) | 44.7 (27.4, 70.7) |
|  | Retired | 52.4 (34.1, 67.2) | 67.8 (47.4, 90.2) | 51.7 (38.8, 72.4) | 45.0 (24.6, 82.4) | 61.1 (43.0, 72.8) | 67.1 (50.9, 82.5) | 47.7 (25.9, 78.5) | 51.2 (34.0, 100) |
|  | Not working/Unemployed | 34.9 (25.3, 54.2) | 48.9 (18.8, 67.0) | 55.4 (42.1, 73.5) | 53.3 (35.4, 73.8) | 44.3 (29.7, 69.9) | 51.3 (24.9, 72.6) | 41.1 (33.7, 66.5) | 40.7 (32.7, 68.4) |
| **Child in house** |  |  |  |  |  |  |  |  |  |
|  | Yes | 38.2 (21.4, 53.5) | 46.5 (32.4, 68.9) | 54.6 (40.3, 68.6) | 55.8 (36.5, 64.7) | 43.5 (32.7, 63.2) | 51.5 (34.2, 63.2) | 39.9 (29.7, 79.4) | 40.5 (30.1, 69.6) |
|  | No | 43.2 (27.9, 58.0) | 56.2 (34.6, 80.4) | 51.5 (42.8, 66.7) | 46.2 (25.0, 66.7) | 53.5 (39.6, 65.1) | 57.8 (41.8, 75.9) | 43.7 (27.9, 65.2) | 44.4 (28.9, 70.0) |
|  | Prefer not to say/Other | 32.5 (22.4, 51.9) | 37.2 (19.1, 78.7) | 56.1 (20.2, 100) | 54.8 (24.5, 100) | 41.7 (24.5, 100) | 49.6 (36.9, 60.4) | 35.1 (19.3, 100) | 47.9 (9.3, 77.1) |
| **Long-term health condition** |  |  |  |  |  |  |  |  |  |
|  | Yes | 47.7 (29.2, 61.9) | 60.0 (40.6, 82.4) | 53.6 (44.6, 69.4) | 45.1 (28.4, 73.7) | 53.0 (39.6, 70.3) | 59.5 (42.6, 77.1) | 47.6 (37.2, 70.9) | 49.1 (35.4, 79.2) |
|  | No | 35.9 (22.6, 50.6) | 47.5 (32.3, 72.4) | 53.7 (40.1, 66.5) | 49.7 (27.3, 64.1) | 45.6 (34.9, 61.6) | 53.1 (36.0, 69.3) | 35.2 (21.3, 65.2) | 35.3 (27.8, 64.5) |
|  | Prefer not to say/Other | 36.4 (15.3, 64.2) | 48.6 (30.1, 76.0) | 51.4 (31.0, 70.1) | 49.9 (34.2, 65.7) | 43.3 (18.0, 74.3) | 51.3 (27.7, 70.1) | 43.7 (21.7, 59.3) | 41.6 (18.1, 62.5) |
| **Frontline worker** |  |  |  |  |  |  |  |  |  |
|  | Yes | 39.6 (22.5, 52.9) | 53.6 (36.9, 80.3) | 53.0 (39.0, 67.4) | 45.9 (23.8, 65.0) | 47.2 (34.2, 65.8) | 56.5 (36.8, 73.9) | 42.2 (24.4, 64.5) | 40.1 (33.4, 64.0) |
|  | No | 41.0 (18.2, 53.9) | 50.3 (33.4, 72.1) | 52.9 (41.0, 66.8) | 51.4 (30.2, 65.0) | 46.9 (33.3, 61.6) | 52.8 (33.4, 66.9) | 38.2 (27.8, 70.2) | 37.5 (31.1, 72.8) |
|  | Prefer not to say/Other | 45.6 (28.8, 59.2) | 56.9 (35.7, 78.6) | 52.5 (41.7, 66.8) | 45.9 (29.6, 68.6) | 53.9 (38.4, 67.5) | 56.1 (42.2, 77.6) | 44.5 (31.4, 65.2) | 47.3 (35.3, 69.9) |

\*Median % (and high/low) for the sample of 15 countries

**Figure 1:**
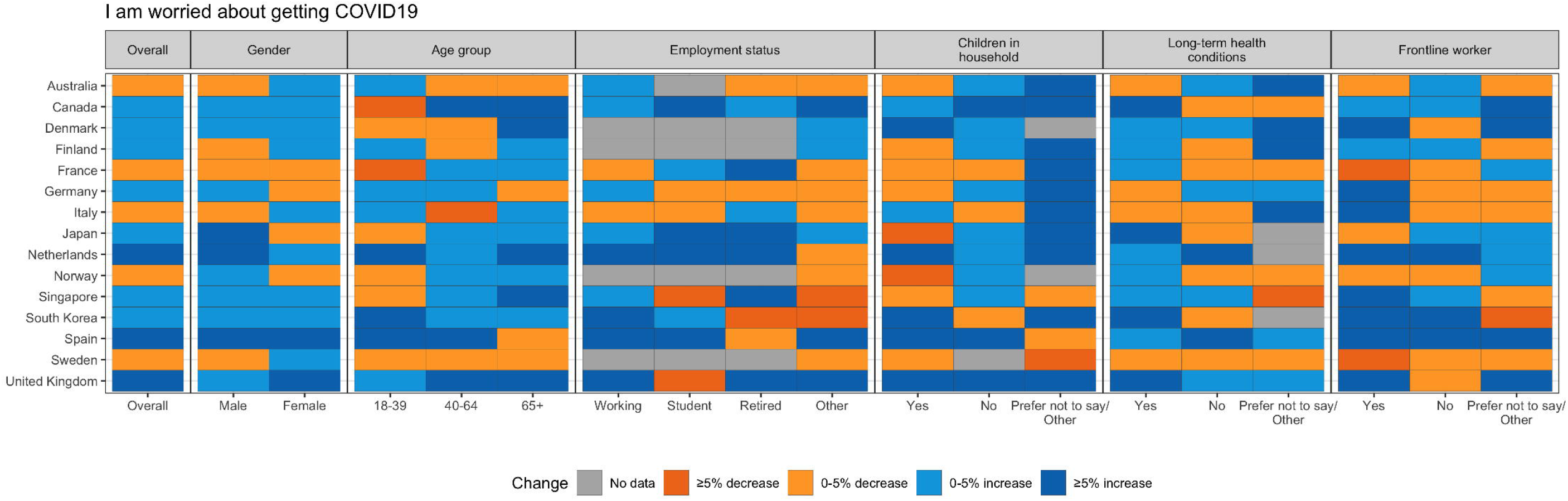
Change in those worried about getting COVID-19.

Across the 15 countries the median estimate for the proportion of respondents that reported they would have the Covid-19 vaccine in the first and second surveys was 40.7% (range: 25.0-55.1) and 55.2% (range: 34.8-77.5), respectively (Table 3). In 12/15 countries there was a significant increase in the proportion of respondents who reported they would have a vaccine (Table 2), for 11/15 countries this increase was at least 5% (Figure 2). In 3/15 countries (Australia, Singapore and South Korea) there was a significant decrease in the proportion that reported they would have the vaccine. Across all population sub-groups, the median estimate for the proportion reporting they would have a vaccine increased between the two surveys (Table 3). On the second survey, the median proportion that reported they would have a vaccine remained below 50% amongst females (49.4%, range: 30.2-79.1), those aged 18-39 years (42.1%, range: 25.9-71.7), those not working or unemployed (48.9, range: 18.8-67.0), students (45.9%, range: 22.8-70.0), those with children at home (46.5%, range: 32.4-68.9) and those without long-term health condition (47.5%, range: 32.3-72.4). This means that for at least half the 15 countries, less than half of those sampled in these groups would have the COVID-19 vaccine.

**Figure 2:**
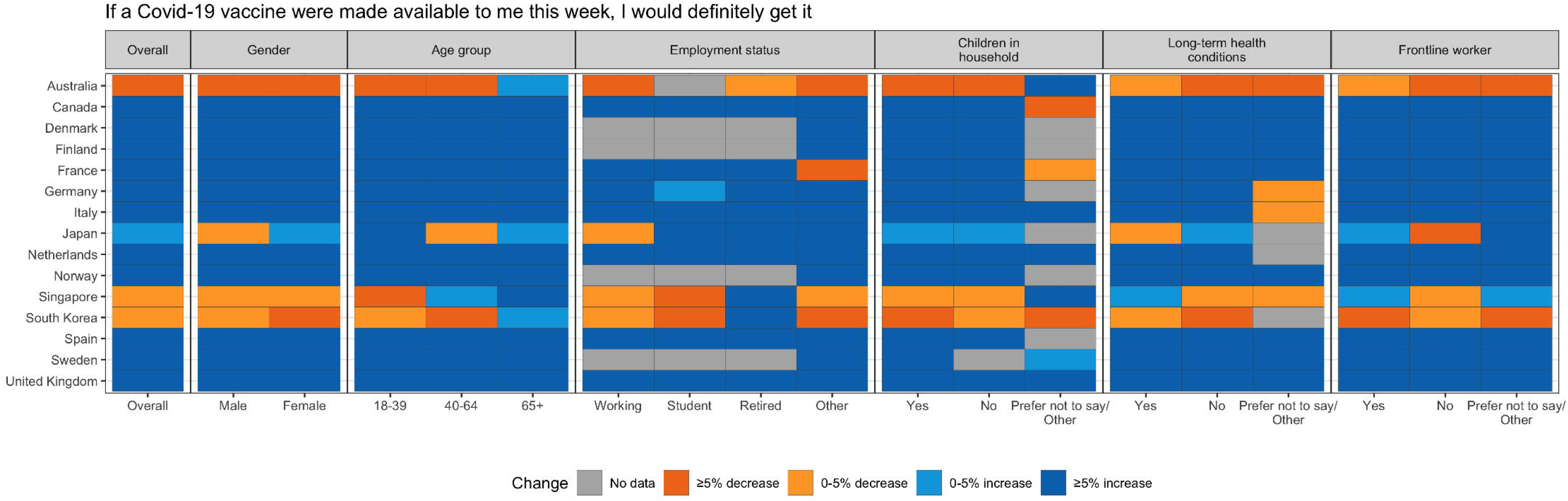
Change in those who ‘agree’ with getting vaccine.

Across the 15 countries the median proportion of respondents that reported they were worried about the side-effects of a COVID-19 vaccine in the first and second surveys was 53.3% (range: 42.1-66.7) and 47.9% (range: 28.0-66.1), respectively (Table 3). In 11/15 countries there was a significant decrease in the proportion of respondents who reported they were worried about the side-effects of a vaccine (Table 2), for 10/15 countries this decrease was at least 5% (Figure 3). In 4/15 countries (Australia, Japan, Singapore and South Korea) there was a significant increase in the proportion that reported they were worried about the side-effects of a vaccine. On the second survey, the median proportion of respondents that reported they were worried about side-effects of a vaccine remained above 50% amongst females (53.7%, range: 31.8-70.4), those aged 18-39 years (50.8%, range: 28.2-60.7), those aged 40-64 years (51.3%, range: 30.7-68.5), those that were working (50.5%, range: 26.7-65.0), those not working or unemployed (53.3, range: 35.4-73.8), those with children at home (55.8%, range: 36.5-64.7) and those that were not frontline workers (51.4%, range: 30.2-65.0). This means that for at least half the 15 countries, more than half of those sampled in these groups were worried about the side-effects of a COVID-19 vaccine.

**Figure 3:**
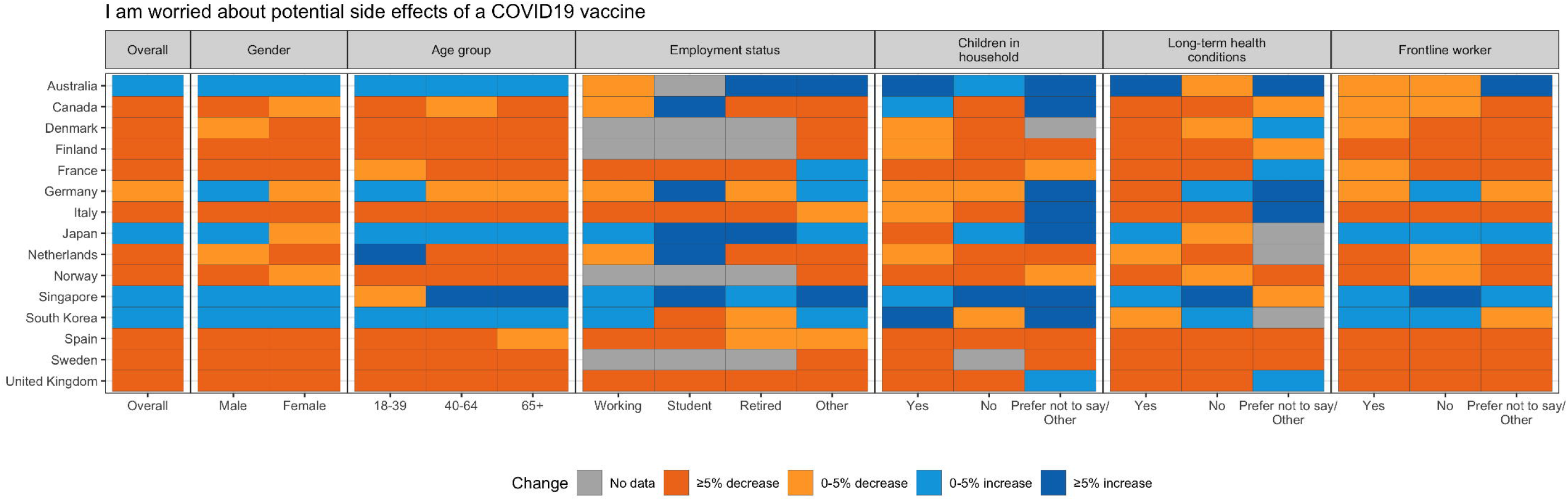
Change in those who ‘agree’ with worried about side-effects of vaccine.

Across the 15 countries the median proportion of respondents that reported they believed their government health authority would provide them the COVID-19 vaccine in the first and second surveys was 49.4% (range: 36.3-64.2) and 55.0% (range: 38.1-72.6), respectively (Table 2). In 12/15 countries there was a significant increase in the proportion of respondents that reported they believed their government health authority would provide a vaccine (Table 2), for 9/15 countries this increase was least 5% (Figure 4). On the second survey, the median proportion that reported they believed their Government would provide the COVID-19 vaccine remained relatively low amongst those aged 18-39 years (49.8%, range: 30.1-64.8).

**Figure 4:**
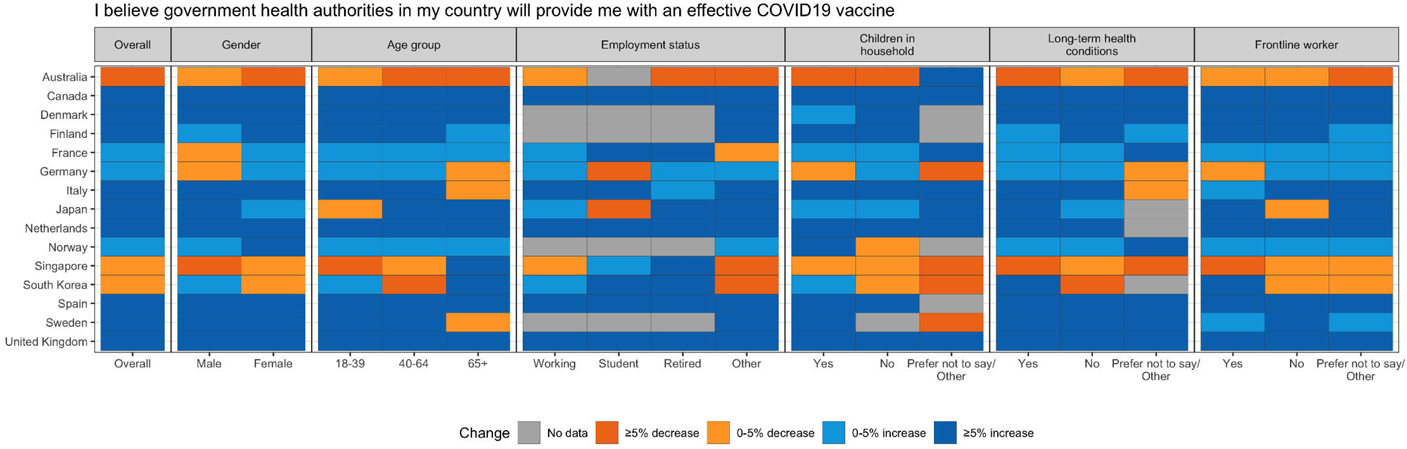
Change in those who ‘agree’ Government health authorities will provide effective vaccine.

## Discussion

This study found willingness to receive a COVID-19 vaccination varied widely across the 15 countries, ranging from 34.8% to 77.5%. However, in all but three of the 15 countries the proportion of respondents who stated they would receive the vaccine increased by more than 5%, over a relatively short time period that coincided with the discovery and roll-out of effective vaccines. In several countries this increase was over 20%. Two of the three countries, where willingness to receive the vaccine decreased, had not started vaccinating their residents at the time the second survey was undertaken.

Previous studies highlight the general public’s concerns around the safety of Covid-19 vaccines^7 8^, we found these concerns remain somewhat high, ranging from 28.0% to 66.1%. It is reassuring that in 11 of the 15 countries surveyed, these concerns have fallen considerably and over a relatively short time. Three of the four countries where respondents reported increased concerns regarding the safety of vaccines, also reported decreased willingness to receive the vaccine. In the majority of countries, respondents reported increased confidence their Governments would provide them with an effective vaccine. Two of the three countries where respondents reported decreased had not yet started their vaccination programmes at the time of the second survey.

An individual’s perception around the safety of a COVID-19 vaccine is a strong predictor to whether they would be willing to receive the vaccine^9^. The public may also be hesitant around the vaccine if they perceive the rapid nature of COVID-19 vaccine research and deployment has been at the expense of adequately exploring their safety. The continued high levels of concern amongst the general population with regards to the safety of COVID-19 vaccines is worrying and may hinder achieving herd immunity. Many countries are already implementing public health messaging that seeks to alleviate these concerns; and these will increasingly need to be targeted at harder to reach populations. Previous studies have found females, younger, and those unemployed to be less willing to receive a COVID-19 vaccine^5 6 9^. This was consistent with our findings. In the majority of the countries, we found willingness to receive the vaccine increased considerably amongst these population groups. However, these three groups remain the most concerned about the side effects of a COVID-19 vaccine. Vaccination programmes in countries should ensure communications emphasis the safety of the vaccine amongst these population groups.

The majority of countries have started vaccinating their population by initially targeting higher risk groups; those aged over 65 years and those with long-term health conditions that places them at higher risk of severe infection. It is reassuring to find high and increasing levels of willingness to receive the vaccine, and low and decreasing levels of concern regarding their safety, amongst these populations. These two groups remain the most concerned about contracting COVID-19 infection. In the majority of countries there was increased concerns about contracting COVID-19 infection between the two survey dates. This is likely to reflect the increasing number of cases seen between the two surveys in many of the countries included in this study. Countries will need to continue monitoring this, especially in relation to people’s perception around the protection offered by the different vaccines and increasing anxieties around new COVID-19 variants emerging.

Students attending higher education institutes will likely need a vaccine considering the high risk of COVID-19 clusters and outbreaks^10^; whilst those who work in the frontline more likely to be exposed. Parents decide whether their child receives the vaccine, and whilst children have not been found to be at increased risk of severe illness or drive transmission^11 12^, vaccination may be needed to prevent school closures. We found relatively low willingness to receive the vaccine and relatively high concerns around their safety in these population groups. Surprisingly these groups also reported lower levels of concern regarding contracting COVID-19 infection. As countries expand the populations being offered the vaccine greater emphasis will need to be taken to ensure parents, younger populations and those that work in the frontline are reached by public health messaging around the need for, and safety in being vaccinated. Evidence-base for effective strategies to tackle vaccine hesitancy is limited and have primarily focussed on childhood vaccines^13 14^. Strategies implemented will need to tackle individual-level factors, but should not ignore the importance of utilising community, social and religious networks to address vaccine hesitancy^15^, and will need to be evaluated.

Strengths of our study include the large number of countries for which data is presented, as well as the exploration of changes in attitudes towards the vaccine and infection over time. Possible limitations include the cross-sectional nature of the study, relatively small numbers surveyed in each country, predominantly high-income European countries surveyed and the use of online survey panels. These do limit the conclusions that can be drawn, especially around the casual relationship between vaccine hesitancy and concerns about their safety.

In conclusion, our findings highlight the willingness of the general public to receive a vaccine is growing over time and the populations subgroups that may need to be targeted with tailored public health messaging around the benefits and safety associated with receiving the COVID-19 vaccine. Although in several countries willingness to vaccinate increased considerable, in half the countries surveyed, in many population sub-groups, less than half would have a vaccine and more than half were worried about their’ side-effects. Further data is needed to understand attitudes towards COVID-19 vaccines from low- and middle-income countries, especially those in South America, Middle East and Africa. Follow-up surveys will need to be undertaken in these and other countries to monitor longer term changes in public attitudes towards the COVID-19 vaccine if the goal of herd immunity is to be achieved.

## Data Availability

All data for YouGov surveys are deidentified and publicly available from: https://github.com/YouGov-Data/covid-19-tracker

## Acknowledgements

We thank YouGov Plc for providing us the data. YouGov are commissioned by the UK Government to undertake the surveys.

## Contributors

RFC, MS, SJ, MSL, HM and AD conceived and designed the study. RFC, MS and NMK did statistical analysis. HM wrote first draft of paper. All authors contributed to data interpretation, revision of paper and contributed to discussion. FRC and HM are the guarantors for this study.

## Ethical approval

Not needed.

## References

1. Randolph HE, Barreiro LB. Herd immunity: understanding COVID-19. Immunity 2020;52(5):737–41.

2. Neumann-Böhme S, Varghese NE, Sabat I, et al. Once we have it, will we use it? A European survey on willingness to be vaccinated against COVID-19: Springer, 2020.

3. Murphy J, Vallières F, Bentall RP, et al. Preparing for a COVID-19 vaccine: Identifying and psychologically profiling those who are vaccine hesitant or resistant in two general population samples. 2020

4. Ward JK, Alleaume C, Peretti-Watel P, et al. The French public’s attitudes to a future COVID-19 vaccine: The politicization of a public health issue. Social science & medicine 2020;265:113414.

5. Malik AA, McFadden SM, Elharake J, et al. Determinants of COVID-19 vaccine acceptance in the US. EClinicalMedicine 2020;26:100495.

6. Murphy J, Vallières F, Bentall RP, et al. Psychological characteristics associated with COVID-19 vaccine hesitancy and resistance in Ireland and the United Kingdom. Nature communications 2021;12(1):1–15.

7. Paul E, Steptoe A, Fancourt D. Attitudes towards vaccines and intention to vaccinate against COVID-19: Implications for public health communications. The Lancet Regional Health-Europe 2020:100012.

8. Thunstrom L, Ashworth M, Finnoff D, et al. Hesitancy towards a COVID-19 vaccine and prospects for herd immunity. Available at SSRN 3593098 2020

9. Karlsson LC, Soveri A, Lewandowsky S, et al. Fearing the disease or the vaccine: The case of COVID-19. Personality and individual differences 2021;172:110590.

10. Wilson E, Donovan CV, Campbell M, et al. Multiple COVID-19 clusters on a university campus—North Carolina, August 2020. Morbidity and Mortality Weekly Report 2020;69(39):1416.

11. Rajmil L. Role of children in the transmission of the COVID-19 pandemic: a rapid scoping review. BMJ Paediatr Open 2020;4(1):e000722. doi: 10.1136/bmjpo-2020-000722 [published Online First: 2020/07/01]

12. de Souza TH, Nadal JA, Nogueira RJ, et al. Clinical manifestations of children with COVID-19: a systematic review. Pediatric pulmonology 2020;55(8):1892–99.

13. Olson O, Berry C, Kumar N. Addressing Parental Vaccine Hesitancy towards Childhood Vaccines in the United States: A Systematic Literature Review of Communication Interventions and Strategies. Vaccines (Basel) 2020;8(4) doi: 10.3390/vaccines8040590 [published Online First: 2020/10/15]

14. Jarrett C, Wilson R, O’Leary M, et al. Strategies for addressing vaccine hesitancy - A systematic review. Vaccine 2015;33(34):4180–90. doi: 10.1016/j.vaccine.2015.04.040 [published Online First: 2015/04/22]

15. Prevention ECfD, Control. Catalogue of interventions addressing vaccine hesitancy: ECDC Stockholm, Germany, 2017.

