## Supplementary File for "International attitudes on COVID-19 vaccination: repeat national cross-sectional surveys across 15 countries"

Appendix A: Australia

Appendix B: Canada

Appendix C: Denmark

Appendix D: Finland

Appendix E: France

Appendix F: Germany

Appendix G: Italy

Appendix H: Japan

Appendix I: Netherlands

Appendix J: Norway

Appendix K: Singapore

Appendix L: South Korea

Appendix M: Spain

Appendix N: Sweden

Appendix O: United Kingdom

**Appendix A: Australia**

| **Variable** | **Category** | **% Would vaccinate** | | | **% Worried about side effects** | | | **% Believe government health authority will provide** | | | **% Worried about getting COVID-19** | | |
| --- | --- | --- | --- | --- | --- | --- | --- | --- | --- | --- | --- | --- | --- |
|  |  | **Wave 1** | **Wave 2** | **Change + 95%CI** | **Wave 1** | **Wave 2** | **Change + 95%CI** | **Wave 1** | **Wave 2** | **Change + 95%CI** | **Wave 1** | **Wave 2** | **Change + 95%CI** |
| **All** | | 55.1 | 48.5 | -6.6 (-6.6, -6.6) | 47.1 | 50.2 | 3.1 (3.0, 3.2) | 60.2 | 55.0 | -5.2 (-5.2, -5.3) | 36.8 | 36.2 | -0.6 (-0.6, -0.6) |
| **Age group** | 18-39 | 49.6 | 43.0 | -6.6 (-6.6, -6.6) | 49.3 | 54.0 | 4.7 (4.6, 4.8) | 55.0 | 52.9 | -2.1 (-2.2, -2.1) | 40.1 | 40.6 | 0.5 (0.4,0.6) |
|  | 40-64 | 55.2 | 44.9 | -10.3 (-10.3,  -10.3) | 50.0 | 51.6 | 1.6 (1.5,1.7) | 57.8 | 52.4 | -5.4 (-5.4, -5.4) | 32.8 | 31.2 | -1.6 (-1.7, -1.6) |
|  | 65+ | 65.8 | 66.8 | 1.0 (0.9,1.1) | 36.9 | 39.4 | 2.5 (2.4,2.6) | 75.5 | 64.2 | -11.3 (-11.3, -11.3) | 38.1 | 36.4 | -1.7 (-1.8, -1.6) |
| **Sex** | Female | 49.9 | 42.5 | -7.4 (-7.4, -7.4) | 52.2 | 56.0 | 3.8 (3.7,3.9) | 58.9 | 53.0 | -5.9 (-5.9, -5.9) | 36.4 | 37.0 | 0.6 (0.5, 0.7) |
|  | Male | 60.5 | 54.7 | -5.8 (-5.8, -5.8) | 41.8 | 44.2 | 2.4 (2.3, 2.5) | 61.6 | 57.0 | -4.6 (-4.6, -4.6) | 37.2 | 35.3 | -1.9 (-1.9, -1.9) |
| **Employment status** | Working | 52.9 | 46.1 | -6.8 (-6.8, -6.8) | 51.3 | 50.2 | -1.1 (-1.2, -1.1) | 57.9 | 55.5 | -2.4 (-2.4, -2.4) | 36.4 | 36.8 | 0.4 (0.3,0.5) |
|  | Student | N/A | N/A |  | N/A | N/A |  | N/A | N/A |  | N/A | N/A |  |
|  | Retired | 65.0 | 64.5 | -0.5 (-0.6, -0.4) | 38.8 | 44.7 | 5.9 (5.7,6.01) | 72.8 | 62.9 | -9.9 (-9.9, -9.9) | 37.8 | 34.5 | -3.3 (-3.4, -3.2) |
|  | Other | 50.5 | 40.4 | -10.1 (-10.1, -10.1) | 44.7 | 55.0 | 10.3 (10.1,10.5) | 53.1 | 46.5 | -6.6 (-6.6, -6.6) | 36.8 | 36.0 | -0.8 (-0.9, -0.7) |
| **Child in house** | No | 55.8 | 50.3 | -5.5 (-5.5, -5.5) | 45.9 | 47.5 | 1.6 (1.5, 1.7) | 62.4 | 57.0 | -5.4 (-5.4, -5.4) | 34.8 | 34.9 | 0.1 (0.1,0.2) |
|  | Prefer not to say/Other | 51.9 | 78.7 | 26.8 (25.8, 27.8) | 50.6 | 80.8 | 30.2 (29.2,31.2) | 24.8 | 40.5 | 15.7 (14.8,16.7) | 27.0 | 38.2 | 11.2 (10.3,12.1) |
|  | Yes | 53.5 | 44.1 | -9.4 (-9.4, -9.4) | 49.7 | 55.8 | 6.1 (6.0, 6.2) | 55.8 | 50.6 | -5.2 (-5.2, -5.2) | 41.5 | 39.1 | -2.4 (-2.5, -2.3) |
| **Long-term health condition** | Yes | 58.1 | 53.9 | -4.2 (-4.2, -4.2) | 44.8 | 50.1 | 5.3 (5.2,5.4) | 63.1 | 56.7 | -6.4 (-6.4, -6.4) | 37.8 | 35.4 | -2.4 (-2.4, -2.4) |
|  | No | 50.6 | 43.1 | -7.5 (-7.5, -7.5) | 49.4 | 49.0 | -0.4 (-0.5, -0.3) | 56.9 | 53.5 | -3.4 (-3.4, -3.4) | 35.0 | 35.3 | 0.3 (0.2,0.4) |
|  | Prefer not to say/Other | 64.2 | 50.0 | -14.2 (-14.3, -14.1) | 53.4 | 65.7 | 12.3 (11.9,12.7) | 57.0 | 52.0 | -5 (-5.2, -4.8) | 42.8 | 55.0 | 12.2 (11.8,12.6) |
| **Frontline worker** | Yes | 50.4 | 48.7 | -1.7 (-1.8, -1.6) | 50.6 | 47.5 | -3.1 (-3.2, -3.0) | 60.8 | 57.1 | -3.7 (-3.8, -3.6) | 41.0 | 38.1 | -2.9 (-3.0, -2.8) |
|  | No | 53.9 | 45.0 | -8.9 (-8.9, -8.9) | 51.6 | 51.4 | -0.2 (-0.3, -0.1) | 56.7 | 54.8 | -1.9 (-2.0, -1.9) | 34.4 | 36.2 | 1.8 (1.7,1.9) |
|  | Prefer not to say/Other | 57.9 | 51.9 | -6 (-6.0, -6.0) | 41.7 | 50.1 | 8.4 (8.3,8.6) | 63.2 | 54.3 | -8.9 (-8.9, -8.9) | 37.3 | 35.3 | -2 (-2.1, -2.0) |

**Appendix B: Canada**

| **Variable** | **Category** | **% Would vaccinate** | | | **% Worried about side effects** | | | **% Believe government health authority will provide** | | | **% Worried about getting COVID-19** | | |
| --- | --- | --- | --- | --- | --- | --- | --- | --- | --- | --- | --- | --- | --- |
|  |  | **Wave 1** | **Wave 2** | **Change + 95%CI** | **Wave 1** | **Wave 2** | **Change + 95%CI** | **Wave 1** | **Wave 2** | **Change + 95%CI** | **Wave 1** | **Wave 2** | **Change + 95%CI** |
| **All** | | 44.2 | 55.2 | 11.0 (10.9,11.2) | 53.3 | 47.9 | -5.4 (-5.4, -5.5) | 49.4 | 58.5 | 9.1 (9.0,9.2) | 38.5 | 42.4 | 3.9 (3.8,4.0) |
| **Age group** | Male | 47.9 | 62.0 | 14.1 (13.9,14.3) | 48.1 | 42.2 | -5.9 (-5.9, -5.9) | 51.3 | 61.3 | 10.0 (9.8,10.2) | 35.4 | 38.7 | 3.3 (3.2,3.4) |
|  | Female | 40.7 | 48.5 | 7.8 (7.7,7.9) | 58.3 | 53.4 | -4.9 (-4.9, -4.9) | 47.5 | 55.8 | 8.3 (8.2,8.5) | 41.5 | 46.1 | 4.6 (4.5,4.7) |
|  | 18-39 | 36.1 | 43.8 | 7.7 (7.6,7.9) | 56.6 | 50.3 | -6.3 (-6.3, -6.3) | 42.7 | 52.7 | 10.0 (9.8,10.2) | 41.5 | 36.1 | -5.4 (-5.4, -5.4) |
| **Sex** | 40-64 | 40.6 | 53.0 | 12.4 (12.2,12.6) | 55.4 | 51.3 | -4.1 (-4.1, -4.1) | 47.3 | 54.1 | 6.8 (6.7,6.9) | 35.9 | 46.2 | 10.3 (10.1,10.5) |
|  | 65+ | 63.3 | 75.8 | 12.5 (12.3,12.7) | 44.3 | 38.1 | -6.2 (-6.2, -6.2) | 63.5 | 75.2 | 11.7 (11.5,11.9) | 38.8 | 44.7 | 5.9 (5.8,6.0) |
| **Employment status** | Working | 39.5 | 51.3 | 11.8 (11.6,12.0) | 54.3 | 50.5 | -3.8 (-3.8, -3.8) | 43.5 | 54.2 | 10.7 (10.5,10.9) | 38.2 | 40.3 | 2.1 (2.0,2.2) |
|  | Student | 52.1 | 61.9 | 9.8 (9.5,10.1) | 39.0 | 47.7 | 8.7 (8.4,9.0) | 58.9 | 72.9 | 14.0 (13.7,14.3) | 41.5 | 46.9 | 5.4 (5.2,5.7) |
|  | Retired | 60.6 | 74.8 | 14.2 (14.0,14.4) | 48.8 | 38.4 | -10.4 (-10.4, -10.4) | 65.4 | 73.7 | 8.3 (8.1,8.5) | 43.2 | 47.0 | 3.8 (3.7,3.9) |
|  | Other | 33.7 | 39.7 | 6.0 (5.9,6.2) | 59.3 | 53.3 | -6.0 (-6.0, -6.0) | 40.6 | 46.8 | 6.2 (6.1,6.4) | 33.7 | 40.3 | 6.6 (6.5,6.8) |
| **Child in house** | Yes | 34.4 | 46.5 | 12.1 (11.9,12.3) | 58.1 | 58.7 | 0.6 (0.5,0.7) | 43.5 | 51.5 | 8 (7.8,8.2) | 39.6 | 40.1 | 0.5 (0.4,0.6) |
|  | No | 50.3 | 62.0 | 11.7 (11.5,11.9) | 52.0 | 42.0 | -10.0 (-10.0, -10.0) | 53.9 | 63.4 | 9.5 (9.4,9.7) | 39.1 | 44.4 | 5.3 (5.2,5.4) |
|  | Prefer not to say/Other | 31.3 | 22.4 | -8.9 (-9.0, -8.8) | 31.8 | 52.7 | 20.9 (20.5,21.3) | 27.5 | 40.1 | 12.6 (12.3,13.0) | 19.3 | 31.1 | 11.8 (11.5,12.1) |
| **Long-term health condition** | Yes | 51.1 | 63.3 | 12.2 (12.0,12.4) | 51.6 | 45.1 | -6.5 (-6.5, -6.5) | 51.9 | 59.5 | 7.6 (7.5,7.7) | 39.0 | 49.0 | 10 (9.8,10.2) |
|  | No | 33.2 | 44.3 | 11.1 (10.9,11.3) | 56.1 | 51.0 | -5.1 (-5.1, -5.1) | 42.2 | 54.6 | 12.4 (12.2,12.6) | 33.0 | 32.2 | -0.8 (-0.9, -0.7) |
|  | Prefer not to say/Other | 46.2 | 53.0 | 6.8 (6.6,7.0) | 52.7 | 49.4 | -3.3 (-3.4, -3.2) | 54.5 | 61.7 | 7.2 (7.0,7.4) | 45.1 | 42.4 | -2.7 (-2.8, -2.6) |
| **Frontline worker** | No | 41.0 | 48.7 | 7.7 (7.5,7.9) | 55.0 | 51.4 | -3.6 (-3.7, -3.6) | 42.6 | 52.8 | 10.2 (10.0,10.4) | 37.3 | 40.4 | 3.1 (3.0,3.2) |
|  | Prefer not to say/Other | 47.5 | 58.2 | 10.7 (10.5,10.9) | 52.6 | 45.9 | -6.7 (-6.7, -6.7) | 53.4 | 61.9 | 8.5 (8.4,8.6) | 38.7 | 44.1 | 5.4 (5.3,5.5) |
|  | Yes | 36.6 | 55.6 | 19 (18.7,19.3) | 53.0 | 49.0 | -4.0 (-4.1, -3.9) | 45.3 | 56.5 | 11.2 (11.0,11.4) | 40.0 | 40.1 | 0.1 (-0.01,0.2) |

**Appendix C: Denmark**

| **Variable** | **Category** | **% Would vaccinate** | | | **% Worried about side effects** | | | **% Believe government health authority will provide** | | | **% Worried about getting COVID-19** | | |
| --- | --- | --- | --- | --- | --- | --- | --- | --- | --- | --- | --- | --- | --- |
|  |  | **Wave 1** | **Wave 2** | **Change + 95%CI** | **Wave 1** | **Wave 2** | **Change + 95%CI** | **Wave 1** | **Wave 2** | **Change + 95%CI** | **Wave 1** | **Wave 2** | **Change + 95%CI** |
| **All** | | 53.7 | 67.0 | 13.3 (13.1,13.5) | 42.1 | 35.4 | -6.7 (-6.7, -6.7) | 63.5 | 72.6 | 9.1 (9.0,9.2) | 37.5 | 39.6 | 2.1 (2.0,2.2) |
| **Age group** | Male | 57.5 | 66.9 | 9.4 (9.2,9.6) | 29.6 | 28.4 | -1.2 (-1.3, -1.2) | 62.4 | 70.8 | 8.4 (8.3,8.5) | 30.7 | 32.8 | 2.1 (2.0,2.2) |
|  | Female | 50.0 | 67.1 | 17.1 (16.9,17.3) | 54.4 | 42.2 | -12.2 (-12.3, -12.1) | 64.5 | 74.3 | 9.8 (9.6, 10.0) | 44.1 | 46.3 | 2.2 (2.1,2.3) |
|  | 18-39 | 36.0 | 50.5 | 14.5 (14.3,14.7) | 42.1 | 37.0 | -5.1 (-5.1, -5.2) | 52.3 | 60.6 | 8.3 (8.1,8.5) | 31.5 | 31.5 | 0 (-0.1,0.1) |
| **Sex** | 40-64 | 58.3 | 68.1 | 9.8 (9.6,10.0) | 44.7 | 36.8 | -7.9 (-7.9, -7.9) | 63.5 | 74.2 | 10.7 (10.5,10.9) | 40.6 | 40.1 | -0.5 (-0.6, -0.4) |
|  | 65+ | 71.9 | 89.5 | 17.6 (17.4,17.9) | 37.3 | 30.5 | -6.8 (-6.8, -6.8) | 80.3 | 87.3 | 7.0 (6.9,7.1) | 40.8 | 50.8 | 10 (9.8,10.2) |
| **Employment status** | Working | N/A | N/A |  | N/A | N/A |  | N/A | N/A |  | N/A | N/A |  |
|  | Student | N/A | N/A |  | N/A | N/A |  | N/A | N/A |  | N/A | N/A |  |
|  | Retired | N/A | N/A |  | N/A | N/A |  | N/A | N/A |  | N/A | N/A |  |
|  | Other | 53.7 | 67.0 | 13.3 (13.1,13.5) | 42.1 | 35.4 | -6.7 (-6.7, -6.7) | 63.5 | 72.6 | 9.1 (9.0,9.2) | 37.5 | 39.6 | 2.1 (2.0,2.2) |
| **Child in house** | Yes | 47.8 | 57.8 | 10 (9.8,10.2) | 40.3 | 39.5 | -0.8 (-0.9, -0.7) | 59.0 | 60.6 | 1.6 (1.5,1.7) | 34.2 | 40.5 | 6.3 (6.1,6.5) |
|  | No | 55.8 | 69.5 | 13.7 (13.5,13.9) | 42.8 | 34.2 | -8.6 (-8.6, -8.6) | 65.1 | 75.9 | 10.8 (10.7,11.0) | 38.7 | 39.3 | 0.6 (0.5,0.7) |
|  | Prefer not to say/Other | N/A | N/A |  | N/A | N/A |  | N/A | N/A |  | N/A | N/A |  |
| **Long-term health condition** | Yes | 61.9 | 71.8 | 9.9 (9.7,10.1) | 44.6 | 34.9 | -9.7 (-9.7, -9.7) | 66.5 | 77.1 | 10.6 (10.4,10.8) | 45.2 | 47.4 | 2.2 (2.1,2.3) |
|  | No | 47.9 | 63.3 | 15.4 (15.2,15.6) | 40.1 | 35.4 | -4.7 (-4.7, -4.7) | 61.6 | 69.0 | 7.4 (7.3,7.5) | 31.8 | 31.9 | 0.1 (0.0,0.2) |
|  | Prefer not to say/Other | 38.0 | 55.2 | 17.2 (16.8,17.7) | 41.8 | 42.1 | 0.3 (0.1,0.6) | 53.0 | 63.2 | 10.2 (9.8,10.6) | 27.1 | 49.9 | 22.8 (22.3,23.3) |
| **Frontline worker** | Yes | 50.2 | 68.7 | 18.5 (18.2,18.8) | 39.0 | 36.0 | -3 (-3.1, -2.9) | 62.5 | 73.9 | 11.4 (11.2,11.6) | 32.0 | 37.6 | 5.6 (5.4,5.8) |
|  | No | 49.8 | 60.0 | 10.2 (10.0,10.4) | 42.9 | 35.3 | -7.6 (-7.6, -7.6) | 60.4 | 66.9 | 6.5 (6.4,6.6) | 35.7 | 32.1 | -3.6 (-3.6, -3.6) |
|  | Prefer not to say/Other | 59.2 | 73.1 | 13.9 (13.7,14.1) | 42.4 | 35.0 | -7.4 (-7.4, -7.4) | 67.3 | 77.6 | 10.3 (10.1,10.5) | 41.4 | 48.5 | 7.1 (7.0,7.2) |

**Appendix D: Finland**

| **Variable** | **Category** | **% Would vaccinate** | | | **% Worried about side effects** | | | **% Believe government health authority will provide** | | | **% Worried about getting COVID-19** | | |
| --- | --- | --- | --- | --- | --- | --- | --- | --- | --- | --- | --- | --- | --- |
|  |  | **Wave 1** | **Wave 2** | **Change + 95%CI** | **Wave 1** | **Wave 2** | **Change + 95%CI** | **Wave 1** | **Wave 2** | **Change + 95%CI** | **Wave 1** | **Wave 2** | **Change + 95%CI** |
| **All** | | 36.6 | 57.0 | 20.4 (20.15,20.65) | 57.3 | 47.2 | -10.1 (-10.16, -10.04) | 47.3 | 53.8 | 6.5 (6.39,6.61) | 34.0 | 35.1 | 1.1 (1.05,1.15) |
| **Age group** | 18-39 | 23.1 | 38.3 | 15.2 (14.98,15.42) | 57.1 | 50.9 | -6.2 (-6.22, -6.18) | 32.3 | 40.5 | 8.2 (8.04,8.36) | 24.8 | 27.8 | 3 (2.9,3.1) |
|  | 40-64 | 35.9 | 60.8 | 24.9 (24.59,25.21) | 59.4 | 49.1 | -10.3 (-10.34, -10.26) | 49.6 | 57.5 | 7.9 (7.75,8.05) | 37.6 | 37.3 | -0.3 (-0.36, -0.24) |
|  | 65+ | 59.3 | 80.5 | 21.2 (20.9,21.5) | 53.3 | 36.3 | -17 (-17.07, -16.93) | 66.2 | 68.3 | 2.1 (1.98,2.22) | 41.0 | 42.6 | 1.6 (1.48,1.72) |
| **Sex** | Female | 29.3 | 58.9 | 29.6 (29.24,29.96) | 69.4 | 55.2 | -14.2 (-14.28, -14.12) | 47.3 | 56.2 | 8.9 (8.75,9.05) | 38.3 | 41.3 | 3 (2.91,3.09) |
|  | Male | 44.2 | 54.9 | 10.7 (10.53,10.87) | 44.8 | 39.0 | -5.8 (-5.81, -5.79) | 47.3 | 51.3 | 4 (3.9,4.1) | 29.5 | 28.7 | -0.8 (-0.85, -0.75) |
| **Employment status** | Working | N/A | N/A |  | N/A | N/A |  | N/A | N/A |  | N/A | N/A |  |
|  | Student | N/A | N/A |  | N/A | N/A |  | N/A | N/A |  | N/A | N/A |  |
|  | Retired | N/A | N/A |  | N/A | N/A |  | N/A | N/A |  | N/A | N/A |  |
|  | Other | 36.6 | 57.0 | 20.4 (20.15,20.65) | 57.3 | 47.2 | -10.1 (-10.16, -10.04) | 47.3 | 53.8 | 6.5 (6.39,6.61) | 34.0 | 35.1 | 1.1 (1.05,1.15) |
| **Child in house** | Yes | 28.4 | 46.0 | 17.6 (17.32,17.88) | 58.9 | 57.6 | -1.3 (-1.39, -1.21) | 40.8 | 48.6 | 7.8 (7.62,7.98) | 32.9 | 30.1 | -2.8 (-2.87, -2.73) |
|  | No | 38.7 | 59.6 | 20.9 (20.64,21.16) | 56.9 | 44.8 | -12.1 (-12.17, -12.03) | 49.1 | 55.2 | 6.1 (5.99,6.21) | 34.3 | 36.2 | 1.9 (1.83,1.97) |
|  | Prefer not to say/Other | N/A | N/A |  | N/A | N/A |  | N/A | N/A |  | N/A | N/A |  |
| **Long-term health condition** | Yes | 44.6 | 66.5 | 21.9 (21.62,22.18) | 55.9 | 45.0 | -10.9 (-10.94, -10.86) | 53.0 | 57.8 | 4.8 (4.69,4.91) | 37.6 | 40.4 | 2.8 (2.71,2.89) |
|  | No | 28.4 | 47.5 | 19.1 (18.85,19.35) | 59.2 | 49.7 | -9.5 (-9.53, -9.47) | 42.0 | 50.3 | 8.3 (8.15,8.45) | 30.3 | 29.3 | -1 (-1.05,-0.95) |
|  | Prefer not to say/Other | 38.4 | 47.1 | 8.7 (8.31,9.09) | 50.1 | 46.2 | -3.9 (-4.16, -3.64) | 38.8 | 42.8 | 4 (3.66,4.34) | 33.9 | 40.5 | 6.6 (6.24,6.96) |
| **Frontline worker** | Yes | 26.3 | 56.8 | 30.5 (30.07,30.93) | 63.1 | 41.4 | -21.7 (-21.78, -21.62) | 34.2 | 54.9 | 20.7 (20.36,21.04) | 28.5 | 33.4 | 4.9 (4.72,5.08) |
|  | No | 27.8 | 51.5 | 23.7 (23.39,24.01) | 58.0 | 52.7 | -5.3 (-5.32, -5.28) | 42.2 | 49.8 | 7.6 (7.45,7.75) | 30.0 | 31.1 | 1.1 (1.02,1.18) |
|  | Prefer not to say/Other | 45.6 | 60.5 | 14.9 (14.69,15.11) | 55.6 | 45.3 | -10.3 (-10.34, -10.26) | 53.9 | 56.1 | 2.2 (2.12,2.28) | 38.2 | 38.1 | -0.1 (-0.16, -0.04) |

**Appendix E: France**

| **Variable** | **Category** | **% Would vaccinate** | | | **% Worried about side effects** | | | **% Believe government health authority will provide** | | | **% Worried about getting COVID-19** | | |
| --- | --- | --- | --- | --- | --- | --- | --- | --- | --- | --- | --- | --- | --- |
|  |  | **Wave 1** | **Wave 2** | **Change + 95%CI** | **Wave 1** | **Wave 2** | **Change + 95%CI** | **Wave 1** | **Wave 2** | **Change + 95%CI** | **Wave 1** | **Wave 2** | **Change + 95%CI** |
| **All** | | 25.0 | 39.2 | 14.2 (14.02,14.38) | 66.2 | 55.2 | -11 (-11.07, -10.93) | 36.3 | 38.1 | 1.8 (1.74,1.86) | 43.1 | 41.8 | -1.3 (-1.33, -1.27) |
| **Age group** | Male | 33.2 | 49.0 | 15.8 (15.58,16.02) | 56.9 | 43.4 | -13.5 (-13.57, -13.43) | 43.2 | 42.7 | -0.5 (-0.56,-0.44) | 40.5 | 38.8 | -1.7 (-1.75, -1.65) |
|  | Female | 17.6 | 30.2 | 12.6 (12.42,12.78) | 74.6 | 65.9 | -8.7 (-8.73, -8.67) | 30.0 | 34.0 | 4 (3.9,4.1) | 45.4 | 44.6 | -0.8 (-0.85, -0.75) |
|  | 18-39 | 19.5 | 25.9 | 6.4 (6.27,6.53) | 63.8 | 60.7 | -3.1 (-3.15, -3.05) | 29.3 | 30.1 | 0.8 (0.72,0.88) | 38.3 | 32.9 | -5.4 (-5.42, -5.38) |
| **Sex** | 40-64 | 23.0 | 40.8 | 17.8 (17.56,18.04) | 67.2 | 56.2 | -11 (-11.05, -10.95) | 37.6 | 39.7 | 2.1 (2.01,2.19) | 43.6 | 44.8 | 1.2 (1.12,1.28) |
|  | 65+ | 36.0 | 56.8 | 20.8 (20.5,21.1) | 67.5 | 44.2 | -23.3 (-23.44, -23.16) | 43.2 | 47.5 | 4.3 (4.16,4.44) | 48.5 | 49.7 | 1.2 (1.09,1.31) |
| **Employment status** | Working | 19.5 | 34.5 | 15 (14.8,15.2) | 65.6 | 57.3 | -8.3 (-8.32, -8.28) | 34.1 | 34.3 | 0.2 (0.14,0.26) | 41.6 | 37.5 | -4.1 (-4.12, -4.08) |
|  | Student | 25.5 | 42.3 | 16.8 (16.46,17.14) | 54.8 | 36.4 | -18.4 (-18.4, -18.4) | 39.0 | 48.6 | 9.6 (9.32,9.88) | 30.2 | 30.7 | 0.5 (0.33,0.67) |
|  | Retired | 34.1 | 59.9 | 25.8 (25.46,26.14) | 65.5 | 45.0 | -20.5 (-20.62, -20.38) | 43.0 | 50.9 | 7.9 (7.73,8.07) | 48.0 | 53.1 | 5.1 (4.96,5.24) |
|  | Other | 25.3 | 18.8 | -6.5 (-6.53, -6.47) | 73.5 | 73.8 | 0.3 (0.2,0.4) | 29.7 | 24.9 | -4.8 (-4.86, -4.74) | 43.2 | 42.6 | -0.6 (-0.71, -0.49) |
| **Child in house** | Yes | 21.4 | 32.4 | 11 (10.83,11.17) | 65.9 | 58.0 | -7.9 (-7.91, -7.89) | 32.7 | 34.2 | 1.5 (1.42,1.58) | 39.5 | 38.3 | -1.2 (-1.25, -1.15) |
|  | No | 27.9 | 46.8 | 18.9 (18.65,19.15) | 66.5 | 52.3 | -14.2 (-14.28, -14.12) | 39.6 | 41.8 | 2.2 (2.12,2.28) | 46.3 | 44.8 | -1.5 (-1.55, -1.45) |
|  | Prefer not to say/Other | 23.8 | 19.1 | -4.7 (-4.95, -4.45) | 64.3 | 59.6 | -4.7 (-5, -4.4) | 27.0 | 42.6 | 15.6 (15.11,16.09) | 34.9 | 57.6 | 22.7 (22.13,23.27) |
| **Long-term health condition** | Yes | 29.2 | 46.7 | 17.5 (17.25,17.75) | 66.1 | 51.2 | -14.9 (-14.98, -14.82) | 39.6 | 42.6 | 3 (2.9,3.1) | 49.5 | 51.8 | 2.3 (2.2,2.4) |
|  | No | 22.6 | 35.0 | 12.4 (12.22,12.58) | 66.5 | 57.4 | -9.1 (-9.14, -9.06) | 34.9 | 36.0 | 1.1 (1.03,1.17) | 39.9 | 37.0 | -2.9 (-2.93, -2.87) |
|  | Prefer not to say/Other | 21.7 | 33.8 | 12.1 (11.67,12.53) | 55.6 | 59.2 | 3.6 (3.23,3.97) | 18.0 | 27.7 | 9.7 (9.31,10.09) | 21.7 | 18.1 | -3.6 (-3.84, -3.36) |
| **Frontline worker** | Yes | 22.5 | 37.8 | 15.3 (15.04,15.56) | 64.1 | 60.4 | -3.7 (-3.78, -3.62) | 36.0 | 36.8 | 0.8 (0.67,0.93) | 46.2 | 40.0 | -6.2 (-6.26, -6.14) |
|  | No | 18.2 | 33.4 | 15.2 (14.98,15.42) | 66.2 | 56.2 | -10 (-10.03, -9.97) | 33.3 | 33.4 | 0.1 (0.03,0.17) | 39.5 | 36.6 | -2.9 (-2.94, -2.86) |
|  | Prefer not to say/Other | 30.4 | 44.2 | 13.8 (13.6,14) | 66.8 | 53.0 | -13.8 (-13.88, -13.72) | 38.4 | 42.2 | 3.8 (3.7,3.9) | 44.5 | 46.5 | 2 (1.92,2.08) |

**Appendix F: Germany**

| **Variable** | **Category** | **% Would vaccinate** | | | **% Worried about side effects** | | | **% Believe government health authority will provide** | | | **% Worried about getting COVID-19** | | |
| --- | --- | --- | --- | --- | --- | --- | --- | --- | --- | --- | --- | --- | --- |
|  |  | **Wave 1** | **Wave 2** | **Change + 95%CI** | **Wave 1** | **Wave 2** | **Change + 95%CI** | **Wave 1** | **Wave 2** | **Change + 95%CI** | **Wave 1** | **Wave 2** | **Change + 95%CI** |
| **All** | | 40.0 | 53.0 | 13 (12.83,13.17) | 51.7 | 50.8 | -0.9 (-0.94, -0.86) | 56.1 | 57.3 | 1.2 (1.14,1.26) | 37.9 | 38.4 | 0.5 (0.45,0.55) |
| **Age group** | Male | 46.5 | 58.6 | 12.1 (11.92,12.28) | 44.6 | 47.8 | 3.2 (3.1,3.3) | 62.7 | 60.5 | -2.2 (-2.24, -2.16) | 37.6 | 39.5 | 1.9 (1.82,1.98) |
|  | Female | 33.8 | 47.7 | 13.9 (13.7,14.1) | 58.4 | 53.7 | -4.7 (-4.72, -4.68) | 49.8 | 54.2 | 4.4 (4.29,4.51) | 38.2 | 37.4 | -0.8 (-0.85, -0.75) |
|  | 18-39 | 31.4 | 42.0 | 10.6 (10.42,10.78) | 45.8 | 49.7 | 3.9 (3.78,4.02) | 52.2 | 52.6 | 0.4 (0.32,0.48) | 33.5 | 34.2 | 0.7 (0.62,0.78) |
| **Sex** | 40-64 | 40.8 | 53.9 | 13.1 (12.91,13.29) | 55.6 | 52.9 | -2.7 (-2.74, -2.66) | 55.0 | 57.0 | 2 (1.92,2.08) | 41.2 | 42.6 | 1.4 (1.32,1.48) |
|  | 65+ | 52.6 | 68.8 | 16.2 (15.93,16.47) | 50.8 | 47.4 | -3.4 (-3.47, -3.33) | 65.9 | 65.5 | -0.4 (-0.5, -0.3) | 36.3 | 35.0 | -1.3 (-1.39, -1.21) |
| **Employment status** | Working | 36.7 | 51.1 | 14.4 (14.2,14.6) | 51.1 | 50.5 | -0.6 (-0.66, -0.54) | 55.5 | 57.1 | 1.6 (1.52,1.68) | 38.0 | 39.7 | 1.7 (1.62,1.78) |
|  | Student | 41.5 | 46.5 | 5 (4.76,5.24) | 46.7 | 54.5 | 7.8 (7.53,8.07) | 60.3 | 51.1 | -9.2 (-9.29, -9.11) | 36.2 | 34.9 | -1.3 (-1.47, -1.13) |
|  | Retired | 51.8 | 67.8 | 16 (15.75,16.25) | 51.7 | 47.0 | -4.7 (-4.75, -4.65) | 64.1 | 65.7 | 1.6 (1.49,1.71) | 40.7 | 40.3 | -0.4 (-0.49, -0.31) |
|  | Other | 34.0 | 39.4 | 5.4 (5.23,5.57) | 55.4 | 56.0 | 0.6 (0.48,0.72) | 45.7 | 47.5 | 1.8 (1.67,1.93) | 34.8 | 32.7 | -2.1 (-2.19, -2.01) |
| **Child in house** | Yes | 30.5 | 40.7 | 10.2 (10,10.4) | 26.8 | 35.5 | 8.7 (7.91,9.49) | 53.1 | 40.0 | -13.1 (-13.76, -12.44) | 27.3 | 39.4 | 12.1 (11.24,12.96) |
|  | No | 42.9 | 55.4 | 12.5 (12.33,12.67) | 51.0 | 50.6 | -0.4 (-0.45, -0.35) | 57.7 | 58.8 | 1.1 (1.04,1.16) | 37.7 | 38.3 | 0.6 (0.54,0.66) |
|  | Prefer not to say/Other | 36.9 | N/A |  | 54.6 | 52.7 | -1.9 (-1.99, -1.81) | 50.4 | 49.7 | -0.7 (-0.8,-0.6) | 39.1 | 39.0 | -0.1 (-0.2,0) |
| **Long-term health condition** | Yes | 45.3 | 60.0 | 14.7 (14.49,14.91) | 55.1 | 49.8 | -5.3 (-5.31, -5.29) | 61.2 | 62.1 | 0.9 (0.83,0.97) | 46.5 | 44.3 | -2.2 (-2.24, -2.16) |
|  | No | 35.0 | 47.9 | 12.9 (12.71,13.09) | 49.4 | 51.9 | 2.5 (2.41,2.59) | 51.5 | 53.5 | 2 (1.91,2.09) | 30.1 | 32.6 | 2.5 (2.41,2.59) |
|  | Prefer not to say/Other | 37.3 | 35.0 | -2.3 (-2.46, -2.14) | 44.2 | 50.3 | 6.1 (5.85,6.35) | 51.9 | 46.9 | -5 (-5.15, -4.85) | 32.7 | 36.4 | 3.7 (3.48,3.92) |
| **Frontline worker** | Yes | 44.4 | 52.5 | 8.1 (7.91,8.29) | 48.9 | 45.9 | -3 (-3.08, -2.92) | 61.5 | 57.3 | -4.2 (-4.27, -4.13) | 37.4 | 43.8 | 6.4 (6.23,6.57) |
|  | No | 33.7 | 50.3 | 16.6 (16.36,16.84) | 51.9 | 53.1 | 1.2 (1.11,1.29) | 53.2 | 57.0 | 3.8 (3.69,3.91) | 38.2 | 37.5 | -0.7 (-0.77,-0.63) |
|  | Prefer not to say/Other | 43.8 | 55.1 | 11.3 (11.12,11.48) | 52.4 | 51.1 | -1.3 (-1.35, -1.25) | 56.7 | 57.4 | 0.7 (0.63,0.77) | 37.9 | 37.0 | -0.9 (-0.95, -0.85) |

**Appendix G: Italy**

| **Variable** | **Category** | **% Would vaccinate** | | | **% Worried about side effects** | | | **% Believe government health authority will provide** | | | **% Worried about getting COVID-19** | | |
| --- | --- | --- | --- | --- | --- | --- | --- | --- | --- | --- | --- | --- | --- |
|  |  | **Wave 1** | **Wave 2** | **Change + 95%CI** | **Wave 1** | **Wave 2** | **Change + 95%CI** | **Wave 1** | **Wave 2** | **Change + 95%CI** | **Wave 1** | **Wave 2** | **Change + 95%CI** |
| **All** | | 40.7 | 56.1 | 15.4 (15.2,15.6) | 48.4 | 40.8 | -7.6 (-7.63, -7.57) | 44.2 | 52.5 | 8.3 (8.17,8.43) | 46.8 | 45.2 | -1.6 (-1.63, -1.57) |
| **Age group** | Male | 46.8 | 58.5 | 11.7 (11.52,11.88) | 42.9 | 34.6 | -8.3 (-8.32, -8.28) | 47.4 | 54.7 | 7.3 (7.16,7.44) | 42.2 | 38.4 | -3.8 (-3.83, -3.77) |
|  | Female | 35.0 | 53.8 | 18.8 (18.55,19.05) | 53.5 | 46.6 | -6.9 (-6.91, -6.89) | 41.2 | 50.5 | 9.3 (9.14,9.46) | 51.1 | 51.6 | 0.5 (0.43,0.57) |
|  | 18-39 | 37.5 | 52.7 | 15.2 (14.96,15.44) | 45.7 | 36.8 | -8.9 (-8.91, -8.89) | 39.6 | 50.4 | 10.8 (10.61,10.99) | 44.0 | 48.1 | 4.1 (3.97,4.23) |
| **Sex** | 40-64 | 38.1 | 55.1 | 17 (16.77,17.23) | 49.8 | 44.0 | -5.8 (-5.8, -5.8) | 41.3 | 51.7 | 10.4 (10.24,10.56) | 48.8 | 42.6 | -6.2 (-6.2, -6.2) |
|  | 65+ | 53.8 | 65.6 | 11.8 (11.57,12.03) | 48.4 | 37.1 | -11.3 (-11.3,-11.3) | 60.8 | 59.4 | -1.4 (-1.5,-1.3) | 45.2 | 49.2 | 4 (3.85,4.15) |
| **Employment status** | Working | 42.2 | 55.4 | 13.2 (13,13.4) | 47.8 | 41.3 | -6.5 (-6.5, -6.5) | 43.4 | 51.8 | 8.4 (8.25,8.55) | 43.7 | 42.7 | -1 (-1.05, -0.95) |
|  | Student | 43.2 | 50.6 | 7.4 (7.15,7.65) | 33.9 | 24.6 | -9.3 (-9.37, -9.23) | 46.1 | 54.7 | 8.6 (8.34,8.86) | 43.3 | 42.3 | -1 (-1.16, -0.84) |
|  | Retired | 52.4 | 68.8 | 16.4 (16.13,16.67) | 48.9 | 34.8 | -14.1 (-14.13, -14.07) | 59.2 | 61.5 | 2.3 (2.17,2.43) | 47.7 | 48.7 | 1 (0.88,1.12) |
|  | Other | 29.1 | 51.1 | 22 (21.69,22.31) | 53.1 | 48.5 | -4.6 (-4.65, -4.55) | 34.8 | 47.9 | 13.1 (12.88,13.32) | 52.7 | 49.9 | -2.8 (-2.86,-2.74) |
| **Child in house** | Yes | 39.1 | 53.3 | 14.2 (13.98,14.42) | 47.8 | 45.2 | -2.6 (-2.65, -2.55) | 39.4 | 47.4 | 8 (7.85,8.15) | 45.5 | 46.2 | 0.7 (0.62,0.78) |
|  | No | 41.5 | 57.7 | 16.2 (15.98,16.42) | 49.5 | 37.9 | -11.6 (-11.66, -11.54) | 46.8 | 55.7 | 8.9 (8.75,9.05) | 47.9 | 44.3 | -3.6 (-3.62, -3.58) |
|  | Prefer not to say/Other | 41.2 | 63.0 | 21.8 (21.2,22.4) | 26.9 | 43.4 | 16.5 (15.96,17.04) | 45.3 | 60.4 | 15.1 (14.57,15.63) | 36.0 | 56.3 | 20.3 (19.72,20.88) |
| **Long-term health condition** | Yes | 46.7 | 59.7 | 13 (12.8,13.2) | 49.9 | 43.6 | -6.3 (-6.31, -6.29) | 52.0 | 57.2 | 5.2 (5.08,5.32) | 52.1 | 52.0 | -0.1 (-0.17, -0.03) |
|  | No | 36.2 | 54.0 | 17.8 (17.56,18.04) | 48.0 | 38.5 | -9.5 (-9.54, -9.46) | 39.1 | 50.0 | 10.9 (10.73,11.07) | 44.0 | 40.9 | -3.1 (-3.13, -3.07) |
|  | Prefer not to say/Other | 45.2 | 43.0 | -2.2 (-2.51, -1.89) | 33.7 | 61.0 | 27.3 (26.67,27.93) | 33.8 | 31.4 | -2.4 (-2.69, -2.11) | 26.2 | 38.4 | 12.2 (11.73,12.67) |
| **Frontline worker** | Yes | 39.6 | 47.8 | 8.2 (7.99,8.41) | 47.7 | 38.6 | -9.1 (-9.13, -9.07) | 46.7 | 51.6 | 4.9 (4.73,5.07) | 39.4 | 45.0 | 5.6 (5.42,5.78) |
|  | No | 43.3 | 57.8 | 14.5 (14.28,14.72) | 47.8 | 42.2 | -5.6 (-5.62, -5.58) | 42.0 | 51.8 | 9.8 (9.63,9.97) | 45.6 | 42.0 | -3.6 (-3.64, -3.56) |
|  | Prefer not to say/Other | 39.3 | 56.9 | 17.6 (17.36,17.84) | 48.9 | 40.2 | -8.7 (-8.72, -8.68) | 44.9 | 53.5 | 8.6 (8.45,8.75) | 49.6 | 48.3 | -1.3 (-1.35, -1.25) |

**Appendix H: Japan**

| **Variable** | **Category** | **% Would vaccinate** | | | **% Worried about side effects** | | | **% Believe government health authority will provide** | | | **% Worried about getting COVID-19** | | |
| --- | --- | --- | --- | --- | --- | --- | --- | --- | --- | --- | --- | --- | --- |
|  |  | **Wave 1** | **Wave 2** | **Change + 95%CI** | **Wave 1** | **Wave 2** | **Change + 95%CI** | **Wave 1** | **Wave 2** | **Change + 95%CI** | **Wave 1** | **Wave 2** | **Change + 95%CI** |
| **All** | | 39.1 | 39.9 | 0.8 (0.73,0.87) | 64.7 | 66.1 | 1.4 (1.33,1.47) | 43.6 | 48.7 | 5.1 (4.99,5.21) | 67.3 | 69.9 | 2.6 (2.51,2.69) |
| **Age group** | Male | 44.0 | 43.9 | -0.1 (-0.19, -0.01) | 57.5 | 61.5 | 4 (3.87,4.13) | 44.3 | 51.0 | 6.7 (6.54,6.86) | 62.4 | 68.6 | 6.2 (6.05,6.35) |
|  | Female | 34.5 | 36.2 | 1.7 (1.6,1.8) | 71.4 | 70.4 | -1 (-1.07, -0.93) | 43.0 | 46.5 | 3.5 (3.38,3.62) | 71.9 | 71.1 | -0.8 (-0.87, -0.73) |
|  | 18-39 | 27.6 | 35.5 | 7.9 (7.7,8.1) | 55.9 | 57.0 | 1.1 (0.96,1.24) | 43.0 | 42.9 | -0.1 (-0.22,0.02) | 62.3 | 61.4 | -0.9 (-1.01, -0.79) |
| **Sex** | 40-64 | 41.1 | 38.6 | -2.5 (-2.56, -2.44) | 66.2 | 68.2 | 2 (1.9,2.1) | 42.1 | 49.3 | 7.2 (7.04,7.36) | 67.3 | 71.6 | 4.3 (4.18,4.42) |
|  | 65+ | 50.4 | 54.4 | 4 (3.79,4.21) | 73.1 | 74.9 | 1.8 (1.63,1.97) | 49.2 | 57.3 | 8.1 (7.85,8.35) | 75.4 | 79.4 | 4 (3.81,4.19) |
| **Employment status** | Working | 43.5 | 39.0 | -4.5 (-4.54, -4.46) | 63.4 | 65.0 | 1.6 (1.51,1.69) | 45.7 | 46.7 | 1 (0.91,1.09) | 68.9 | 69.9 | 1 (0.92,1.08) |
|  | Student | 28.5 | 45.3 | 16.8 (16.3,17.3) | 33.4 | 40.7 | 7.3 (6.89,7.71) | 66.9 | 45.4 | -21.5 (-21.62, -21.38) | 38.0 | 54.5 | 16.5 (16,17) |
|  | Retired | 55.5 | 70.5 | 15 (14.54,15.46) | 72.4 | 82.4 | 10 (9.63,10.37) | 50.2 | 76.4 | 26.2 (25.64,26.76) | 75.2 | 100.0 | 24.8 (24.37,25.23) |
|  | Other | 28.8 | 36.9 | 8.1 (7.9,8.3) | 69.2 | 72.1 | 2.9 (2.75,3.05) | 35.2 | 51.3 | 16.1 (15.81,16.39) | 66.5 | 68.4 | 1.9 (1.76,2.04) |
| **Child in house** | Yes | 43.6 | 43.6 | 0 (-0.14,0.14) | 20.2 | 67.2 | 47 (46,48) | 40.5 | 56.1 | 15.6 (14.75,16.45) | 20.2 | 67.2 | 47 (46,48) |
|  | No | 38.6 | 39.4 | 0.8 (0.72,0.88) | 64.3 | 66.7 | 2.4 (2.31,2.49) | 41.0 | 45.7 | 4.7 (4.58,4.82) | 65.2 | 70.0 | 4.8 (4.68,4.92) |
|  | Prefer not to say/Other | 22.5 | NA |  | 68.6 | 63.4 | -5.2 (-5.29, -5.11) | 55.4 | 60.4 | 5 (4.8,5.2) | 79.4 | 69.6 | -9.8 (-9.84, -9.76) |
| **Long-term health condition** | Yes | 47.7 | 46.5 | -1.2 (-1.29, -1.11) | 69.4 | 73.7 | 4.3 (4.16,4.44) | 45.0 | 56.3 | 11.3 (11.08,11.52) | 70.9 | 79.2 | 8.3 (8.12,8.48) |
|  | No | 33.8 | 36.1 | 2.3 (2.2,2.4) | 61.8 | 61.6 | -0.2 (-0.28, -0.12) | 42.7 | 44.3 | 1.6 (1.5,1.7) | 65.2 | 64.5 | -0.7 (-0.77, -0.63) |
|  | Prefer not to say/Other | N/A | N/A |  | N/A | N/A |  | N/A | N/A |  | N/A | N/A |  |
| **Frontline worker** | Yes | 34.3 | 37.5 | 3.2 (2.99,3.41) | 62.9 | 65.0 | 2.1 (1.9,2.3) | 40.5 | 46.8 | 6.3 (6.06,6.54) | 64.5 | 59.8 | -4.7 (-4.83, -4.57) |
|  | No | 46.0 | 39.4 | -6.6 (-6.62, -6.58) | 63.6 | 65.0 | 1.4 (1.3,1.5) | 47.2 | 46.7 | -0.5 (-0.59, -0.41) | 70.2 | 72.8 | 2.6 (2.49,2.71) |
|  | Prefer not to say/Other | 33.2 | 42.0 | 8.8 (8.61,8.99) | 66.3 | 68.6 | 2.3 (2.17,2.43) | 40.7 | 53.3 | 12.6 (12.37,12.83) | 65.2 | 69.9 | 4.7 (4.55,4.85) |

**Appendix I: Netherlands**

| **Variable** | **Category** | **% Would vaccinate** | | | **% Worried about side effects** | | | **% Believe government health authority will provide** | | | **% Worried about getting COVID-19** | | |
| --- | --- | --- | --- | --- | --- | --- | --- | --- | --- | --- | --- | --- | --- |
|  |  | **Wave 1** | **Wave 2** | **Change + 95%CI** | **Wave 1** | **Wave 2** | **Change + 95%CI** | **Wave 1** | **Wave 2** | **Change + 95%CI** | **Wave 1** | **Wave 2** | **Change + 95%CI** |
| **All** | | 44.6 | 63.1 | 18.5 (18.25,18.75) | 46.6 | 41.3 | -5.3 (-5.31, -5.29) | 47.3 | 56.6 | 9.3 (9.14,9.46) | 28.7 | 34.6 | 5.9 (5.78,6.02) |
| **Age group** | Male | 49.0 | 67.0 | 18 (17.73,18.27) | 40.1 | 37.3 | -2.8 (-2.86, -2.74) | 49.2 | 56.5 | 7.3 (7.14,7.46) | 23.8 | 33.7 | 9.9 (9.72,10.08) |
|  | Female | 40.3 | 59.3 | 19 (18.72,19.28) | 52.8 | 45.2 | -7.6 (-7.61, -7.59) | 45.5 | 56.7 | 11.2 (11,11.4) | 33.4 | 35.5 | 2.1 (1.99,2.21) |
|  | 18-39 | 31.9 | 45.0 | 13.1 (12.86,13.34) | 45.5 | 50.8 | 5.3 (5.14,5.46) | 33.5 | 42.9 | 9.4 (9.2,9.6) | 27.8 | 33.3 | 5.5 (5.34,5.66) |
| **Sex** | 40-64 | 44.9 | 67.6 | 22.7 (22.38,23.02) | 48.2 | 37.5 | -10.7 (-10.71, -10.69) | 50.5 | 58.1 | 7.6 (7.43,7.77) | 31.3 | 34.6 | 3.3 (3.18,3.42) |
|  | 65+ | 64.0 | 84.9 | 20.9 (20.56,21.24) | 45.1 | 33.8 | -11.3 (-11.33, -11.27) | 62.7 | 78.7 | 16 (15.71,16.29) | 24.7 | 37.0 | 12.3 (12.04,12.56) |
| **Employment status** | Working | 41.7 | 57.4 | 15.7 (15.46,15.94) | 42.4 | 38.4 | -4 (-4.04, -3.96) | 42.9 | 52.7 | 9.8 (9.62,9.98) | 26.7 | 33.9 | 7.2 (7.05,7.35) |
|  | Student | 38.9 | 64.8 | 25.9 (25.27,26.53) | 59.0 | 70.7 | 11.7 (11.22,12.18) | 40.7 | 53.0 | 12.3 (11.8,12.8) | 27.2 | 52.7 | 25.5 (24.88,26.12) |
|  | Retired | 60.6 | 82.5 | 21.9 (21.54,22.26) | 49.1 | 36.1 | -13 (-13.02, -12.98) | 65.2 | 75.7 | 10.5 (10.25,10.75) | 25.9 | 34.0 | 8.1 (7.87,8.33) |
|  | Other | 38.0 | 63.8 | 25.8 (25.39,26.21) | 54.8 | 49.6 | -5.2 (-5.3, -5.1) | 43.7 | 53.4 | 9.7 (9.45,9.95) | 38.5 | 34.5 | -4 (-4.11, -3.89) |
| **Child in house** | Yes | 38.2 | 61.4 | 23.2 (22.87,23.53) | 45.8 | 45.6 | -0.2 (-0.29, -0.11) | 41.8 | 55.4 | 13.6 (13.37,13.83) | 29.7 | 41.6 | 11.9 (11.69,12.11) |
|  | No | 50.4 | 64.7 | 14.3 (14.07,14.53) | 46.6 | 37.3 | -9.3 (-9.31, -9.29) | 52.1 | 57.6 | 5.5 (5.36,5.64) | 27.9 | 28.9 | 1 (0.91,1.09) |
|  | Prefer not to say/Other | 27.5 | 54.1 | 26.6 (25.78,27.42) | 73.5 | 56.9 | -16.6 (-16.98, -16.22) | 41.7 | 54.1 | 12.4 (11.69,13.11) | 27.2 | 38.0 | 10.8 (10.15,11.45) |
| **Long-term health condition** | Yes | 52.4 | 66.2 | 13.8 (13.57,14.03) | 47.7 | 44.9 | -2.8 (-2.87, -2.73) | 49.3 | 58.7 | 9.4 (9.21,9.59) | 37.2 | 41.8 | 4.6 (4.46,4.74) |
|  | No | 37.8 | 60.1 | 22.3 (21.99,22.61) | 45.6 | 37.9 | -7.7 (-7.71, -7.69) | 45.6 | 54.6 | 9 (8.82,9.18) | 21.3 | 27.8 | 6.5 (6.36,6.64) |
|  | Prefer not to say/Other | N/A | N/A |  | N/A | N/A |  | N/A | N/A |  | N/A | N/A |  |
| **Frontline worker** | Yes | 42.3 | 65.0 | 22.7 (22.32,23.08) | 45.4 | 33.8 | -11.6 (-11.64, -11.56) | 47.2 | 63.9 | 16.7 (16.38,17.02) | 24.4 | 34.6 | 10.2 (9.95,10.45) |
|  | No | 41.4 | 54.2 | 12.8 (12.57,13.03) | 41.0 | 40.3 | -0.7 (-0.79, -0.61) | 41.0 | 48.0 | 7 (6.83,7.17) | 27.8 | 33.5 | 5.7 (5.55,5.85) |
|  | Prefer not to say/Other | 48.7 | 71.6 | 22.9 (22.57,23.23) | 52.5 | 45.7 | -6.8 (-6.83, -6.77) | 53.5 | 62.5 | 9 (8.81,9.19) | 31.4 | 35.7 | 4.3 (4.16,4.44) |

**Appendix J: Norway**

| **Variable** | **Category** | **% Would vaccinate** | | | **% Worried about side effects** | | | **% Believe government health authority will provide** | | | **% Worried about getting COVID-19** | | |
| --- | --- | --- | --- | --- | --- | --- | --- | --- | --- | --- | --- | --- | --- |
|  |  | **Wave 1** | **Wave 2** | **Change + 95%CI** | **Wave 1** | **Wave 2** | **Change + 95%CI** | **Wave 1** | **Wave 2** | **Change + 95%CI** | **Wave 1** | **Wave 2** | **Change + 95%CI** |
| **All** | | 41.9 | 56.5 | 14.6 (14.41,14.79) | 49.9 | 44.0 | -5.9 (-5.91, -5.89) | 54.1 | 57.0 | 2.9 (2.83,2.97) | 41.1 | 40.7 | -0.4 (-0.44, -0.36) |
| **Age group** | Male | 46.5 | 57.9 | 11.4 (11.22,11.58) | 44.5 | 37.9 | -6.6 (-6.6, -6.6) | 55.3 | 55.9 | 0.6 (0.53,0.67) | 35.8 | 37.8 | 2 (1.92,2.08) |
|  | Female | 37.3 | 55.1 | 17.8 (17.56,18.04) | 55.2 | 50.3 | -4.9 (-4.91, -4.89) | 53.0 | 58.1 | 5.1 (4.99,5.21) | 46.4 | 43.7 | -2.7 (-2.74, -2.66) |
|  | 18-39 | 33.6 | 45.4 | 11.8 (11.61,11.99) | 49.6 | 43.6 | -6 (-6.01, -5.99) | 47.7 | 49.8 | 2.1 (2,2.2) | 36.8 | 32.1 | -4.7 (-4.72, -4.68) |
| **Sex** | 40-64 | 41.6 | 56.4 | 14.8 (14.58,15.02) | 50.7 | 44.7 | -6 (-6.01, -5.99) | 52.0 | 55.0 | 3 (2.9,3.1) | 39.4 | 41.8 | 2.4 (2.31,2.49) |
|  | 65+ | 58.0 | 77.0 | 19 (18.72,19.28) | 48.6 | 43.4 | -5.2 (-5.25, -5.15) | 70.4 | 74.3 | 3.9 (3.77,4.03) | 52.6 | 54.4 | 1.8 (1.68,1.92) |
| **Employment status** | Working | N/A | N/A |  | N/A | N/A |  | N/A | N/A |  | N/A | N/A |  |
|  | Student | N/A | N/A |  | N/A | N/A |  | N/A | N/A |  | N/A | N/A |  |
|  | Retired | N/A | N/A |  | N/A | N/A |  | N/A | N/A |  | N/A | N/A |  |
|  | Other | 41.9 | 56.5 | 14.6 (14.41,14.79) | 49.9 | 44.0 | -5.9 (-5.91, -5.89) | 54.1 | 57.0 | 2.9 (2.83,2.97) | 41.1 | 40.7 | -0.4 (-0.44, -0.36) |
| **Child in house** | Yes | 37.9 | 54.9 | 17 (16.74,17.26) | 100.0 | 100.0 | 0 (0,0) | 42.1 | 51.5 | 9.4 (9.21,9.59) | 39.2 | 32.7 | -6.5 (-6.52, -6.48) |
|  | No | 43.5 | 57.0 | 13.5 (13.31,13.69) | 50.4 | 44.9 | -5.5 (-5.5, -5.5) | 58.7 | 58.5 | -0.2 (-0.25, -0.15) | 41.9 | 43.1 | 1.2 (1.14,1.26) |
|  | Prefer not to say/Other | N/A | N/A |  | 48.3 | 40.8 | -7.5 (-7.52, -7.48) | 100.0 | N/A |  |  |  |  |
| **Long-term health condition** | Yes | 48.7 | 58.9 | 10.2 (10.03,10.37) | 51.3 | 44.4 | -6.9 (-6.9, -6.9) | 59.0 | 60.1 | 1.1 (1.02,1.18) | 47.6 | 49.1 | 1.5 (1.42,1.58) |
|  | No | 36.7 | 53.8 | 17.1 (16.87,17.33) | 48.9 | 44.1 | -4.8 (-4.81, -4.79) | 50.5 | 53.1 | 2.6 (2.51,2.69) | 35.2 | 33.4 | -1.8 (-1.84, -1.76) |
|  | Prefer not to say/Other | 35.5 | 61.6 | 26.1 (25.62,26.58) | 46.1 | 40.1 | -6 (-6.16, -5.84) | 47.8 | 68.6 | 20.8 (20.37,21.23) | 44.5 | 39.7 | -4.8 (-4.97, -4.63) |
| **Frontline worker** | Yes | 38.9 | 63.9 | 25 (24.64,25.36) | 49.1 | 35.6 | -13.5 (-13.52, -13.48) | 57.3 | 59.2 | 1.9 (1.77,2.03) | 42.9 | 39.7 | -3.2 (-3.28, -3.12) |
|  | No | 38.1 | 50.0 | 11.9 (11.71,12.09) | 48.8 | 47.2 | -1.6 (-1.66, -1.54) | 49.6 | 51.2 | 1.6 (1.51,1.69) | 36.7 | 33.1 | -3.6 (-3.63, -3.57) |
|  | Prefer not to say/Other | 46.5 | 58.8 | 12.3 (12.11,12.49) | 51.1 | 44.7 | -6.4 (-6.4,-6.4) | 57.3 | 60.7 | 3.4 (3.3,3.5) | 44.6 | 47.3 | 2.7 (2.61,2.79) |

**Appendix K: Singapore**

| **Variable** | **Category** | **% Would vaccinate** | | | **% Worried about side effects** | | | **% Believe government health authority will provide** | | | **% Worried about getting COVID-19** | | |
| --- | --- | --- | --- | --- | --- | --- | --- | --- | --- | --- | --- | --- | --- |
|  |  | **Wave 1** | **Wave 2** | **Change + 95%CI** | **Wave 1** | **Wave 2** | **Change + 95%CI** | **Wave 1** | **Wave 2** | **Change + 95%CI** | **Wave 1** | **Wave 2** | **Change + 95%CI** |
| **All** | | 36.1 | 34.8 | -1.3 (-1.33, -1.27) | 58.9 | 63.6 | 4.7 (4.61,4.79) | 64.2 | 60.3 | -3.9 (-3.9, -3.9) | 47.8 | 49.2 | 1.4 (1.34,1.46) |
| **Age group** | Male | 40.9 | 39.2 | -1.7 (-1.75, -1.65) | 54.9 | 59.4 | 4.5 (4.39,4.61) | 63.7 | 58.2 | -5.5 (-5.51, -5.49) | 47.2 | 49.0 | 1.8 (1.72,1.88) |
|  | Female | 31.5 | 30.5 | -1 (-1.05, -0.95) | 62.7 | 67.6 | 4.9 (4.79,5.01) | 64.6 | 62.3 | -2.3 (-2.34, -2.26) | 48.4 | 49.5 | 1.1 (1.03,1.17) |
|  | 18-39 | 37.6 | 29.6 | -8 (-8.01, -7.99) | 59.5 | 58.1 | -1.4 (-1.46, -1.34) | 65.6 | 58.4 | -7.2 (-7.2, -7.2) | 46.7 | 44.3 | -2.4 (-2.45, -2.35) |
| **Sex** | 40-64 | 34.3 | 35.6 | 1.3 (1.23,1.37) | 59.8 | 68.5 | 8.7 (8.55,8.85) | 63.5 | 59.6 | -3.9 (-3.92, -3.88) | 49.3 | 51.9 | 2.6 (2.51,2.69) |
|  | 65+ | 39.7 | 50.6 | 10.9 (10.65,11.15) | 51.8 | 60.3 | 8.5 (8.27,8.73) | 62.2 | 71.4 | 9.2 (8.97,9.43) | 44.3 | 55.3 | 11 (10.74,11.26) |
| **Employment status** | Working | 37.0 | 34.4 | -2.6 (-2.62, -2.58) | 58.6 | 63.5 | 4.9 (4.8,5) | 62.9 | 58.2 | -4.7 (-4.71, -4.69) | 46.3 | 49.7 | 3.4 (3.31,3.49) |
|  | Student | 36.6 | 31.0 | -5.6 (-5.73, -5.47) | 59.9 | 66.2 | 6.3 (6.05,6.55) | 70.8 | 73.7 | 2.9 (2.7,3.1) | 53.7 | 42.4 | -11.3 (-11.38, -11.22) |
|  | Retired | 34.4 | 47.4 | 13 (12.71,13.29) | 56.4 | 57.3 | 0.9 (0.74,1.06) | 61.1 | 75.4 | 14.3 (14.01,14.59) | 45.1 | 51.2 | 6.1 (5.88,6.32) |
|  | Other | 32.3 | 29.6 | -2.7 (-2.79, -2.61) | 61.6 | 67.3 | 5.7 (5.52,5.88) | 69.9 | 54.7 | -15.2 (-15.23, -15.17) | 54.8 | 48.9 | -5.9 (-5.97, -5.83) |
| **Child in house** | Yes | 39.5 | 35.0 | -4.5 (-4.53, -4.47) | 61.9 | 64.7 | 2.8 (2.69,2.91) | 63.2 | 58.5 | -4.7 (-4.73, -4.67) | 51.0 | 49.5 | -1.5 (-1.56, -1.44) |
|  | No | 34.9 | 34.6 | -0.3 (-0.35, -0.25) | 57.4 | 62.9 | 5.5 (5.39,5.61) | 64.8 | 61.7 | -3.1 (-3.12,-3.08) | 45.8 | 49.0 | 3.2 (3.11,3.29) |
|  | Prefer not to say/Other | 24.4 | 36.3 | 11.9 (11.52,12.28) | 56.9 | 66.7 | 9.8 (9.43,10.17) | 61.9 | 49.6 | -12.3 (-12.45, -12.15) | 53.9 | 52.0 | -1.9 (-2.15, -1.65) |
| **Long-term health condition** | Yes | 36.7 | 40.6 | 3.9 (3.78,4.02) | 58.8 | 62.8 | 4 (3.88,4.12) | 70.3 | 65.2 | -5.1 (-5.13, -5.07) | 50.8 | 51.5 | 0.7 (0.61,0.79) |
|  | No | 35.9 | 32.3 | -3.6 (-3.62, -3.58) | 58.8 | 64.1 | 5.3 (5.19,5.41) | 60.9 | 57.9 | -3 (-3.02, -2.98) | 46.3 | 48.5 | 2.2 (2.12,2.28) |
|  | Prefer not to say/Other | 33.4 | 30.1 | -3.3 (-3.55, -3.05) | 61.2 | 60.6 | -0.6 (-0.86, -0.34) | 74.3 | 64.2 | -10.1 (-10.28, -9.92) | 51.9 | 40.7 | -11.2 (-11.39, -11.01) |
| **Frontline worker** | Yes | 34.9 | 36.9 | 2 (1.88,2.12) | 60.8 | 64.4 | 3.6 (3.47,3.73) | 65.8 | 59.1 | -6.7 (-6.73, -6.67) | 44.4 | 55.1 | 10.7 (10.49,10.91) |
|  | No | 37.9 | 33.5 | -4.4 (-4.42, -4.38) | 57.6 | 63.2 | 5.6 (5.48,5.72) | 61.6 | 57.9 | -3.7 (-3.72, -3.68) | 47.1 | 47.6 | 0.5 (0.43,0.57) |
|  | Prefer not to say/Other | 33.8 | 35.7 | 1.9 (1.8, 2.0) | 59.7 | 63.8 | 4.1 (3.97,4.23) | 67.5 | 65.7 | -1.8 (-1.86, -1.74) | 51.6 | 48.1 | -3.5 (-3.55, -3.45) |

**Appendix L: South Korea**

| **Variable** | **Category** | **% Would vaccinate** | | | **% Worried about side effects** | | | **% Believe government health authority will provide** | | | **% Worried about getting COVID-19** | | |
| --- | --- | --- | --- | --- | --- | --- | --- | --- | --- | --- | --- | --- | --- |
|  |  | **Wave 1** | **Wave 2** | **Change + 95%CI** | **Wave 1** | **Wave 2** | **Change + 95%CI** | **Wave 1** | **Wave 2** | **Change + 95%CI** | **Wave 1** | **Wave 2** | **Change + 95%CI** |
| **All** | | 48.7 | 43.7 | -5 (-5.01, -4.99) | 55.7 | 57.6 | 1.9 (1.82,1.98) | 54.7 | 53.6 | -1.1 (-1.15, -1.05) | 55.1 | 58.3 | 3.2 (3.1,3.3) |
| **Age group** | Male | 51.1 | 50.4 | -0.7 (-0.79, -0.61) | 49.8 | 53.2 | 3.4 (3.27,3.53) | 54.7 | 56.4 | 1.7 (1.59,1.81) | 53.0 | 56.6 | 3.6 (3.47,3.73) |
|  | Female | 46.4 | 37.1 | -9.3 (-9.3, -9.3) | 61.6 | 62.0 | 0.4 (0.31,0.49) | 54.7 | 50.8 | -3.9 (-3.95, -3.85) | 57.2 | 60.0 | 2.8 (2.68,2.92) |
|  | 18-39 | 40.9 | 37.1 | -3.8 (-3.87, -3.73) | 56.7 | 59.2 | 2.5 (2.37,2.63) | 48.4 | 48.7 | 0.3 (0.19,0.41) | 52.3 | 58.9 | 6.6 (6.42,6.78) |
| **Sex** | 40-64 | 53.5 | 46.6 | -6.9 (-6.92, -6.88) | 53.9 | 54.8 | 0.9 (0.8,1) | 59.1 | 53.5 | -5.6 (-5.63, -5.57) | 55.4 | 56.0 | 0.6 (0.51,0.69) |
|  | 65+ | 49.9 | 51.6 | 1.7 (1.46,1.94) | 64.7 | 66.8 | 2.1 (1.86,2.34) | 52.0 | 70.8 | 18.8 (18.39,19.21) | 65.4 | 67.8 | 2.4 (2.16,2.64) |
| **Employment status** | Working | 49.1 | 44.9 | -4.2 (-4.24, -4.16) | 53.3 | 56.8 | 3.5 (3.39,3.61) | 54.0 | 54.1 | 0.1 (0.02,0.18) | 52.0 | 59.3 | 7.3 (7.15,7.45) |
|  | Student | 41.5 | 22.8 | -18.7 (-18.84, -18.56) | 59.8 | 50.9 | -8.9 (-9.16, -8.64) | 47.6 | 56.2 | 8.6 (8.17,9.03) | 50.4 | 54.9 | 4.5 (4.11,4.89) |
|  | Retired | 45.0 | 61.1 | 16.1 (15.69,16.51) | 65.0 | 60.4 | -4.6 (-4.8, -4.4) | 55.9 | 64.3 | 8.4 (8.06,8.74) | 75.4 | 65.2 | -10.2 (-10.33, -10.07) |
|  | Other | 50.2 | 35.6 | -14.6 (-14.6, -14.6) | 59.2 | 61.4 | 2.2 (2.03,2.37) | 57.9 | 45.9 | -12 (-12.03, -11.97) | 58.7 | 51.7 | -7 (-7.08, -6.92) |
| **Child in house** | Yes | 51.8 | 44.8 | -7 (-7.03, -6.97) | 49.7 | 60.0 | 10.3 (10.1,10.5) | 53.2 | 56.8 | 3.6 (3.46,3.74) | 49.3 | 57.4 | 8.1 (7.92,8.28) |
|  | No | 46.3 | 43.2 | -3.1 (-3.16, -3.04) | 60.5 | 55.5 | -5 (-5.04, -4.96) | 55.7 | 51.3 | -4.4 (-4.44, -4.36) | 60.0 | 58.2 | -1.8 (-1.87, -1.73) |
|  | Prefer not to say/Other | 50.2 | 36.9 | -13.3 (-13.67, -12.93) | 50.9 | 68.7 | 17.8 (17.12,18.48) | 59.7 | 52.7 | -7 (-7.42, -6.58) | 41.5 | 77.1 | 35.6 (34.77,36.43) |
| **Long-term health condition** | Yes | 52.8 | 51.5 | -1.3 (-1.4, -1.2) | 57.7 | 57.5 | -0.2 (-0.3, -0.1) | 54.6 | 68.6 | 14 (13.75,14.25) | 52.7 | 62.5 | 9.8 (9.59,10.01) |
|  | No | 46.4 | 39.4 | -7 (-7.01, -6.99) | 54.6 | 57.7 | 3.1 (2.99,3.21) | 54.7 | 45.2 | -9.5 (-9.51, -9.49) | 56.5 | 55.9 | -0.6 (-0.68, -0.52) |
|  | Prefer not to say/Other | N/A | N/A |  | N/A | N/A |  | N/A | N/A |  | N/A | N/A |  |
| **Frontline worker** | Yes | 52.0 | 42.4 | -9.6 (-9.65, -9.55) | 54.3 | 56.6 | 2.3 (2.13,2.47) | 48.1 | 54.2 | 6.1 (5.89,6.31) | 51.1 | 57.1 | 6 (5.8,6.2) |
|  | No | 47.9 | 46.2 | -1.7 (-1.78, -1.62) | 52.9 | 56.9 | 4 (3.86,4.14) | 56.5 | 54.1 | -2.4 (-2.47, -2.33) | 52.4 | 60.4 | 8 (7.83,8.17) |
|  | Prefer not to say/Other | 48.0 | 40.8 | -7.2 (-7.25, -7.15) | 60.5 | 59.7 | -0.8 (-0.91,-0.69) | 56.1 | 52.3 | -3.8 (-3.88, -3.72) | 61.0 | 55.8 | -5.2 (-5.26, -5.14) |

**Appendix M: Spain**

| **Variable** | **Category** | **% Would vaccinate** | | | **% Worried about side effects** | | | **% Believe government health authority will provide** | | | **% Worried about getting COVID-19** | | |
| --- | --- | --- | --- | --- | --- | --- | --- | --- | --- | --- | --- | --- | --- |
|  |  | **Wave 1** | **Wave 2** | **Change + 95%CI** | **Wave 1** | **Wave 2** | **Change + 95%CI** | **Wave 1** | **Wave 2** | **Change + 95%CI** | **Wave 1** | **Wave 2** | **Change + 95%CI** |
| **All** | | 28.0 | 52.1 | 24.1 (23.82,24.38) | 66.7 | 57.4 | -9.3 (-9.35, -9.25) | 41.0 | 54.8 | 13.8 (13.62,13.98) | 60.1 | 66.0 | 5.9 (5.8,6) |
| **Age group** | Male | 32.8 | 55.0 | 22.2 (21.91,22.49) | 59.3 | 49.3 | -10 (-10.04, -9.96) | 43.4 | 55.9 | 12.5 (12.31,12.69) | 56.8 | 63.1 | 6.3 (6.17,6.43) |
|  | Female | 23.5 | 49.4 | 25.9 (25.58,26.22) | 73.6 | 65.0 | -8.6 (-8.63, -8.57) | 38.7 | 53.9 | 15.2 (14.99,15.41) | 63.2 | 68.8 | 5.6 (5.48,5.72) |
|  | 18-39 | 25.0 | 42.1 | 17.1 (16.86,17.34) | 66.4 | 56.2 | -10.2 (-10.22, -10.18) | 32.5 | 49.0 | 16.5 (16.26,16.74) | 54.1 | 59.5 | 5.4 (5.27,5.53) |
| **Sex** | 40-64 | 28.4 | 57.0 | 28.6 (28.26,28.94) | 67.8 | 57.5 | -10.3 (-10.34, -10.26) | 44.4 | 57.0 | 12.6 (12.41,12.79) | 60.7 | 69.0 | 8.3 (8.16,8.44) |
|  | 65+ | 34.8 | 59.0 | 24.2 (23.82,24.58) | 61.8 | 60.6 | -1.2 (-1.32, -1.08) | 47.7 | 61.7 | 14 (13.72,14.28) | 73.9 | 71.4 | -2.5 (-2.6,-2.4) |
| **Employment status** | Working | 27.3 | 52.6 | 25.3 (24.99,25.61) | 66.9 | 53.9 | -13 (-13.07, -12.93) | 40.3 | 54.6 | 14.3 (14.1,14.5) | 57.2 | 63.4 | 6.2 (6.08,6.32) |
|  | Student | 26.1 | 44.1 | 18 (17.65,18.35) | 53.4 | 48.2 | -5.2 (-5.33, -5.07) | 35.3 | 54.7 | 19.4 (19.03,19.77) | 45.1 | 70.7 | 25.6 (25.17,26.03) |
|  | Retired | 38.8 | 66.0 | 27.2 (26.79,27.61) | 65.0 | 60.7 | -4.3 (-4.39, -4.21) | 54.4 | 67.1 | 12.7 (12.44,12.96) | 78.5 | 76.6 | -1.9 (-2.0, -1.8) |
|  | Other | 25.3 | 48.9 | 23.6 (23.29,23.91) | 69.6 | 64.7 | -4.9 (-4.93, -4.87) | 37.9 | 51.4 | 13.5 (13.28,13.72) | 60.6 | 66.0 | 5.4 (5.27,5.53) |
| **Child in house** | Yes | 27.3 | 48.7 | 21.4 (21.12,21.68) | 74.4 | 24.5 | -49.9 (-50.27, -49.53) | 39.5 | 50.4 | 10.9 (10.72,11.08) | 50.9 | 49.9 | -1 (-1.7, -0.3) |
|  | No | 28.5 | 54.5 | 26 (25.69,26.31) | 66.7 | 56.9 | -9.8 (-9.84, -9.76) | 42.0 | 58.0 | 16 (15.78,16.22) | 59.5 | 65.3 | 5.8 (5.69,5.91) |
|  | Prefer not to say/Other | 50.9 | N/A |  | 66.5 | 58.5 | -8 (-8.01, -7.99) | 24.5 | N/A |  | 61.3 | 67.3 | 6 (5.87,6.13) |
| **Long-term health condition** | Yes | 33.1 | 60.6 | 27.5 (27.16,27.84) | 67.7 | 57.4 | -10.3 (-10.34, -10.26) | 45.4 | 59.6 | 14.2 (13.99,14.41) | 67.9 | 71.3 | 3.4 (3.3,3.5) |
|  | No | 24.5 | 46.4 | 21.9 (21.62,22.18) | 66.0 | 57.5 | -8.5 (-8.53, -8.47) | 37.8 | 51.7 | 13.9 (13.7,14.1) | 54.1 | 62.0 | 7.9 (7.76,8.04) |
|  | Prefer not to say/Other | 15.3 | 34.9 | 19.6 (19.15,20.05) | 64.3 | 56.9 | -7.4 (-7.63, -7.17) | 35.3 | 42.9 | 7.6 (7.22,7.98) | 59.3 | 62.5 | 3.2 (2.87,3.53) |
| **Frontline worker** | Yes | 26.7 | 53.6 | 26.9 (26.51,27.29) | 67.4 | 59.9 | -7.5 (-7.55, -7.45) | 41.4 | 56.0 | 14.6 (14.33,14.87) | 55.7 | 64.0 | 8.3 (8.09,8.51) |
|  | No | 27.5 | 52.3 | 24.8 (24.48,25.12) | 66.8 | 52.0 | -14.8 (-14.88, -14.72) | 40.0 | 54.1 | 14.1 (13.89,14.31) | 57.7 | 63.2 | 5.5 (5.37,5.63) |
|  | Prefer not to say/Other | 28.8 | 51.6 | 22.8 (22.51,23.09) | 66.4 | 61.2 | -5.2 (-5.21, -5.19) | 41.7 | 55.2 | 13.5 (13.3,13.7) | 63.1 | 68.9 | 5.8 (5.68,5.92) |

**Appendix N: Sweden**

| **Variable** | **Category** | **% Would vaccinate** | | | **% Worried about side effects** | | | **% Believe government health authority will provide** | | | **% Worried about getting COVID-19** | | |
| --- | --- | --- | --- | --- | --- | --- | --- | --- | --- | --- | --- | --- | --- |
|  |  | **Wave 1** | **Wave 2** | **Change + 95%CI** | **Wave 1** | **Wave 2** | **Change + 95%CI** | **Wave 1** | **Wave 2** | **Change + 95%CI** | **Wave 1** | **Wave 2** | **Change + 95%CI** |
| **All** | | 34.9 | 57.6 | 22.7 (22.43,22.97) | 54.4 | 41.2 | -13.2 (-13.29, -13.11) | 43.0 | 50.6 | 7.6 (7.48,7.72) | 40.0 | 38.3 | -1.7 (-1.73, -1.67) |
| **Age group** | 18-39 | 25.1 | 41.7 | 16.6 (16.36,16.84) | 50.0 | 40.3 | -9.7 (-9.72, -9.68) | 33.6 | 43.1 | 9.5 (9.33,9.67) | 33.3 | 31.9 | -1.4 (-1.46, -1.34) |
|  | 40-64 | 35.1 | 59.5 | 24.4 (24.09,24.71) | 58.6 | 44.6 | -14 (-14.07, -13.93) | 42.0 | 49.8 | 7.8 (7.65,7.95) | 42.0 | 40.8 | -1.2 (-1.26, -1.14) |
|  | 65+ | 54.8 | 77.2 | 22.4 (22.08,22.72) | 51.8 | 37.0 | -14.8 (-14.85, -14.75) | 65.0 | 62.4 | -2.6 (-2.67, -2.53) | 48.3 | 43.7 | -4.6 (-4.66, -4.54) |
| **Sex** | Female | 27.8 | 59.6 | 31.8 (31.42,32.18) | 63.5 | 46.3 | -17.2 (-17.31, -17.09) | 43.6 | 51.5 | 7.9 (7.76,8.04) | 42.9 | 43.2 | 0.3 (0.23,0.37) |
|  | Male | 42.0 | 55.5 | 13.5 (13.3,13.7) | 45.2 | 36.1 | -9.1 (-9.13, -9.07) | 42.3 | 49.6 | 7.3 (7.16,7.44) | 37.2 | 33.4 | -3.8 (-3.82, -3.78) |
| **Employment status** | Working | N/A | N/A |  | N/A | N/A |  | N/A | N/A |  | N/A | N/A |  |
|  | Student | N/A | N/A |  | N/A | N/A |  | N/A | N/A |  | N/A | N/A |  |
|  | Retired | N/A | N/A |  | N/A | N/A |  | N/A | N/A |  | N/A | N/A |  |
|  | Other | 34.9 | 57.6 | 22.7 (22.43,22.97) | 54.4 | 41.2 | -13.2 (-13.29, -13.11) | 43.0 | 50.6 | 7.6 (7.48,7.72) | 40.0 | 38.3 | -1.7 (-1.73, -1.67) |
| **Child in house** | Yes | 34.9 | 57.8 | 22.9 (22.63,23.17) | 54.3 | 41.4 | -12.9 (-12.99, -12.81) | 42.9 | 50.7 | 7.8 (7.68,7.92) | 39.9 | 38.6 | -1.3 (-1.33, -1.27) |
|  | No | N/A | N/A |  | N/A | N/A |  | N/A | N/A |  | N/A | N/A |  |
|  | Prefer not to say/Other | 33.7 | 37.2 | 3.5 (3,4) | 56.1 | 27.0 | -29.1 (-29.34, -28.86) | 55.3 | 36.9 | -18.4 (-18.76, -18.04) | 55.8 | 9.3 | -46.5 (-46.51, -46.49) |
| **Long-term health condition** | Yes | 44.4 | 63.6 | 19.2 (18.94,19.46) | 53.6 | 40.1 | -13.5 (-13.57, -13.43) | 46.1 | 53.0 | 6.9 (6.76,7.04) | 47.1 | 44.1 | -3 (-3.04, -2.96) |
|  | No | 27.4 | 52.3 | 24.9 (24.59,25.21) | 53.7 | 42.2 | -11.5 (-11.55, -11.45) | 40.7 | 48.3 | 7.6 (7.46,7.74) | 33.0 | 32.4 | -0.6 (-0.65, -0.55) |
|  | Prefer not to say/Other | 26.9 | 52.7 | 25.8 (25.29,26.31) | 70.1 | 41.4 | -28.7 (-28.74, -28.66) | 37.8 | 50.5 | 12.7 (12.32,13.08) | 49.5 | 47.8 | -1.7 (-1.93, -1.47) |
| **Frontline worker** | Yes | 40.3 | 60.2 | 19.9 (19.6,20.2) | 58.6 | 40.9 | -17.7 (-17.77, -17.63) | 45.9 | 50.5 | 4.6 (4.45,4.75) | 45.1 | 38.2 | -6.9 (-6.94, -6.86) |
|  | No | 28.5 | 51.9 | 23.4 (23.1,23.7) | 54.7 | 41.1 | -13.6 (-13.66, -13.54) | 36.1 | 47.9 | 11.8 (11.61,11.99) | 34.7 | 33.3 | -1.4 (-1.45, -1.35) |
|  | Prefer not to say/Other | 39.6 | 61.1 | 21.5 (21.22,21.78) | 52.1 | 41.4 | -10.7 (-10.74, -10.66) | 49.3 | 52.8 | 3.5 (3.4,3.6) | 43.6 | 42.5 | -1.1 (-1.16, -1.04) |

**Appendix O: United Kingdom**

| **Variable** | **Category** | **% Would vaccinate** | | | **% Worried about side effects** | | | **% Believe government health authority will provide** | | | **% Worried about getting COVID-19** | | |
| --- | --- | --- | --- | --- | --- | --- | --- | --- | --- | --- | --- | --- | --- |
|  |  | **Wave 1** | **Wave 2** | **Change + 95%CI** | **Wave 1** | **Wave 2** | **Change + 95%CI** | **Wave 1** | **Wave 2** | **Change + 95%CI** | **Wave 1** | **Wave 2** | **Change + 95%CI** |
| **All** | | 54.3 | 77.5 | 23.2 (22.9,23.5) | 46.3 | 28.0 | -18.3 (-18.4, -18.2) | 50.7 | 70.5 | 19.8 (19.6,20.0) | 44.2 | 50.1 | 5.9 (5.8,6) |
| **Age group** | Male | 59.7 | 75.9 | 16.2 (16.0,16.4) | 40.1 | 24.1 | -16 (-16.1, -15.9) | 54.2 | 69.4 | 15.2 (15.0,15.4) | 40.6 | 44.9 | 4.3 (4.2,4.4) |
|  | Female | 49.1 | 79.1 | 30 (29.6,30.4) | 52.3 | 31.8 | -20.5 (-20.6, -20.4) | 47.3 | 71.5 | 24.2 (23.9,24.5) | 47.7 | 55.0 | 7.3 (7.2,7.4) |
|  | 18-39 | 47.6 | 71.7 | 24.1 (23.8,24.4) | 47.4 | 28.1 | -19.3 (-19.4, -19.2) | 47.2 | 64.8 | 17.6 (17.4,17.9) | 40.1 | 42.4 | 2.3 (2.2,2.4) |
| **Sex** | 40-64 | 52.2 | 76.0 | 23.8 (23.5,24.1) | 47.2 | 30.7 | -16.5 (-16.6, -16.4) | 48.1 | 69.2 | 21.1 (20.8,21.4) | 44.5 | 53.2 | 8.7 (8.5,8.9) |
|  | 65+ | 69.6 | 90.0 | 20.4 (20.1,20.7) | 42.9 | 23.4 | -19.5 (-19.6, -19.4) | 61.4 | 82.2 | 20.8 (20.5,21.1) | 50.9 | 57.9 | 7 (6.8,7.2) |
| **Employment status** | Working | 50.7 | 76.7 | 26 (25.7,26.3) | 47.5 | 26.7 | -20.8 (-21.0, -20.7) | 47.8 | 69.7 | 21.9 (21.6,22.2) | 40.9 | 46.6 | 5.7 (5.6,5.8) |
|  | Student | 38.6 | 70.0 | 31.4 (30.9,31.9) | 47.1 | 13.1 | -34 (-34.2, -33.9) | 54.6 | 74.3 | 19.7 (19.3,20.1) | 40.6 | 27.4 | -13.2 (-13.3, -13.1) |
|  | Retired | 67.2 | 90.2 | 23 (22.7,23.3) | 43.3 | 24.6 | -18.7 (-18.8, -18.6) | 62.0 | 82.5 | 20.5 (20.2,20.8) | 50.9 | 58.6 | 7.7 (7.5,7.9) |
|  | Other | 54.2 | 64.3 | 10.1 (9.9,10.3) | 46.1 | 40.7 | -5.4 (-5.5, -5.3) | 44.3 | 55.1 | 10.8 (10.6,11.0) | 47.7 | 55.1 | 7.4 (7.2,7.6) |
| **Child in house** | Yes | 44.1 | 68.9 | 24.8 (24.5,25.1) | 54.8 | 36.5 | -18.3 (-18.4, -18.2) | 44.1 | 63.2 | 19.1 (18.8,19.4) | 41.1 | 47.4 | 6.3 (6.1,6.5) |
|  | No | 58.0 | 80.4 | 22.4 (22.1,22.7) | 43.8 | 25.0 | -18.8 (-18.9, -18.7) | 53.1 | 73.3 | 20.2 (20.0,20.5) | 45.5 | 51.2 | 5.7 (5.6,5.8) |
|  | Prefer not to say/Other | 22.4 | 75.3 | 52.9 (52.0,53.8) | 34.1 | 38.6 | 4.5 (4.0, 5.0) | 30.8 | 50.3 | 19.5 (18.9,20.1) | 31.0 | 38.6 | 7.6 (7.1,8.1) |
| **Long-term health condition** | Yes | 60.3 | 82.4 | 22.1 (21.8,22.4) | 46.2 | 28.4 | -17.8 (-17.9, -17.7) | 53.9 | 71.6 | 17.7 (17.5,17.9) | 51.6 | 59.0 | 7.4 (7.2,7.5) |
|  | No | 49.1 | 72.4 | 23.3 (23.0,23.6) | 47.5 | 27.3 | -20.2 (-20.3, -20.1) | 48.2 | 69.3 | 21.1 (20.8,21.4) | 36.3 | 40.6 | 4.3 (4.2,4.4) |
|  | Prefer not to say/Other | 34.9 | 76.0 | 41.1 (40.4,41.8) | 31.0 | 34.2 | 3.2 (2.9, 3.5) | 35.7 | 70.1 | 34.4 (33.8,35.0) | 47.6 | 51.1 | 3.5 (3.2,3.8) |
| **Frontline worker** | Yes | 52.9 | 80.3 | 27.4 (27.1,27.8) | 45.1 | 23.8 | -21.3 (-21.4, -21.2) | 48.9 | 72.4 | 23.5 (23.2,23.8) | 42.2 | 53.1 | 10.9 (10.7,11.1) |
|  | No | 48.7 | 72.1 | 23.4 (23.1,23.7) | 49.7 | 30.2 | -19.5 (-19.61, -19.4) | 46.9 | 66.5 | 19.6 (19.3,19.9) | 39.7 | 38.5 | -1.2 (-1.3, -1.1) |
|  | Prefer not to say/Other | 58.6 | 78.6 | 20 (19.7,20.3) | 44.9 | 29.6 | -15.3 (-15.4, -15.2) | 54.2 | 71.4 | 17.2 (17.0,17.4) | 48.4 | 54.4 | 6 (5.9,6.1) |
